# Localized and Whole-Room Effects of Portable Air Filtration Units on Aerosol Particle Deposition and Concentration in a Classroom Environment

**DOI:** 10.1101/2021.08.22.21262392

**Authors:** Meng Kong, Linhao Li, Stephanie M. Eilts, Li Li, Christopher J. Hogan, Zachary C. Pope

## Abstract

In indoor environments with limited ventilation, recirculating portable air filtration (PAF) units may reduce COVID-19 infection risk via not only the direct aerosol route (*i.e.*, inhalation) but also via an indirect aerosol route (*i.e.*, contact with the surface where particles deposited). We systematically investigated the impact of PAF units in a mock classroom, as a supplement to background ventilation, on localized and whole-room surface deposition and particle concentration. Fluorescently tagged particles with a volumetric mean diameter near two micrometers were continuously introduced into the classroom environment via a breathing simulator with a prescibed inhalation-exhalation waveform. Deposition velocities were inferred on >50 horizontal and vertical surfaces throughout the classroom, while aerosol concentrations were spatially monitored via optical particle spectrometry. Results revealed a particle decay rate consistent with expectations based upon the reported clean air delivery rates of the PAF units. Additionally, the PAF units reduced peak concentrations by a factor of around 2.5 compared to the highest concentrations observed and led to a statistically significant reduction in deposition velocities for horizontal surfaces >2.5 m from the aerosol source. Our results not only confirm PAF units can reduce particle concentrations but also demonstrate that they may lead to reduced particle deposition throughout an indoor environment when properly positioned.

**Practical Implications:**

- Portable air filtration units should be prioritized in classrooms as part of a multi-layed strategy to mitigate potentially infectious particle transmission by direct aerosol transmission via inhalation and indirect aerosol transmission via particle deposition to surfaces and later contact with said surfaces.
- When placing portable air filtration unit(s) within a classrom space, one should consider the airflow field within the classroom, the characteristic operational mode (heating vs. cooling) of the heating, ventilation, and air conditioning system, the predominantly occupied areas of the classroom, and interference with the regular teaching and learning activities.

## 1. Introduction

Coronavirus disease-2019 (COVID-19), the infectious disease caused by the severe acute respiratory syndrome coronavirus-2 (SARS-CoV-2),^1, 2^ has led to >207 million confirmed cases and 4.4 million deaths globally^3, 4^ as of August 2021 (the time of writing this manuscript). Like other respiratory infections, the SARS-CoV-2 virus is transmitted via the expulsion of respiratory droplets and particles during breathing, speaking, and other human respiratory activities. The fate of such droplets ultimately determines the mode of exposure for individuals. Human respiratory activities produce an extremely broad and often multimodal size distribution of droplets,^5–11^ but the smallest mode of the size distribution is peaked below 10 micrometers for breathing and speaking, the most common human respiratory activities. Recent studies have repeatedly detected viral RNA in such sub-10 micrometer droplets,^12–16^ strongly suggesting they are largely responsible for infectious virus dispersion into the environment and, ultimately, that they are the primary drivers of infection spread. Droplets in this size range evaporate upon dispersion into indoor environments,^17, 18^ yielding micrometer scale aerosol particles with lifetimes in the air which are tens of minutes-to-hours.^19^ Such particles are easily dispersed throughout the entire indoor space where they originate and can contribute to infection spread via two means. First, a direct aerosol transmission route occurs when infectious particles are inhaled and deposit within the airways. Second, exhaled micrometer scale particles can deposit on surfaces,^19–22^ and readily do so on surfaces throughout indoor spaces. They can then contribute to viral loading through human contact with these surfaces, which we term an indirect aerosol transmission route. While the direct aerosol route has been most prominently discussed in the context of SARS-CoV-2 and may be the primary route of transmission in many instances^23, 24^, it is important to note the indirect route may contribute to viral loading as well, in certain circumstances. Even with perfect deposition efficiency in human airways, exposure levels through direct inhalation are bounded by the minute volume (5-8 L min^-1^). Deposition velocities for microparticles in indoor spaces are often of order 10^-5^-10^-4^ m s^-^^1, 25^ and approximating human surface area as 1.7 m^2^ leads to a deposition exposure equivalent of ∼1.0-10 L min^-1^, similar in magnitude to direct inhalation.

Reducing the potential for infection spread via both direct inhalation and through deposition and contact hence requires limiting expelled aerosol particle concentrations and lifetimes in the environment. Alongside the universal use of highly efficient masks (N95 respirators or equivalent) as both source control and protective equipment, this is best accomplished through increasing ventilation rates, limiting occupancy in rooms, and limiting time of co-occupancy. Limiting occupancy and co-occupancy time is a particular challenge in many settings, notably for K-12 school environments. In the United States, fifty million students and five million adult staff comprise the kindergarten through grade 12 (K-12) public school system.^26, 27^ To limit the spread of COVID-19 among staff and students, many U.S. schools enacted distance learning throughout most of 2020 and into 2021.^28^ However, many school systems are set to open or have opened to in-person instruction in Fall 2021. Infection spread in school systems is hence a persistant concern,^29–31^ and employing interventions that limit respiratory virus transmission is paramount.^32^

Recommendations for classrooms state that proper ventilation should be higher than 3-6 air changes per hour (ACH),^33, 34^ with recent literature highlighting lower COVID-19 infection rates can be achieved in schools when ventilation and/or filtration are improved as part of a multi- level COVID-19 mitigation strategy.^35, 36^ However, not all school systems will be able to achieve such ACH values; many schools have older infrastructure and limited heating, cooling, and air conditioning (HVAC) systems that are unable to adequately ventilate the rooms with fresh air or filter the air being delivered to the space. Recirculating portable air filtration (PAF) units, which typically incorporate fibrous filters to remove particles from air, but may also incorporate other technologies (*i.e.*, to remove [volatile organic compounds] VOCs or to augment filter collection),^37–39^ are a demonstrated approach to supplement ventilation systems.^40, 41^ For this reason, many school systems are considering, or have committed to adding, PAFs to their classrooms. While prior studies have examined PAF performance in mock classroom settings,^40, 42^ they have largely been limited to examining rates of particle concentration decay following a simulated aerosol event, akin to tests used in evaluating clean air delivery rates (CADR) for such units.^43^ Here, we seek to expand on these studies by examining microparticle dispersion from a breathing simulator in a mock classroom (herafter “classroom”) environment having its own closed air handling unit (AHU). Not only do we evaluate the effect of PAFs on aerosol clearance rates, but we also report on the effect they have on deposition velocity^21, 25, 44–46^ on surfaces throughout the classroom. In doing so, we examine the influence PAFs have on the potential for both direct and indirect infection transmission. Specifically, as described in the subsequent sections, we injected 1-3 µm fluorescein-tagged particles via a breathing simulator situated in the classroom space. We evaluated particle deposition fluxes and velocities, as well as concentration, at different locations around the classroom. Four experimental conditions were implemented, with one condition run with HVAC system operation only and the three other conditions with HVAC system operation supplemented by three PAF units running at predefined settings. Each measurement location was a varied distance from the simulator and the three PAF units situated throughout the space, allowing us to examine the localized and whole-room impact of portable air filtration on particle deposition and concentration. The results demonstrate how PAFs can be used to not only reduce airborne particle concentrations, but also reduce deposition velocities, hence directly mitigating both direct and indirect aerosol transmission routes.

## 2. Methods

### 2.1. Lab Configuration & HVAC System

We conducted the experiments in an 88.3 m^2^ area, 229.6 m^3^ volume module (12.8 m length, 6.9 m width, 2.6 m height) within the Well Living Lab in Rochester, MN, shown schematically in Figure 1a. This module was originally configured as an open floorplan office, but was reconfigured to a classroom for 12 students and one teacher. Twelve common school desks were brought into the classroom, each 0.71 m in height. We also placed a teacher’s desk of 0.79 m in height at the northeast corner of the classroom, with a desktop computer and two monitors placed atop at 1.30 m in height. A 1.38 m tall whiteboard was placed at the north end of the classroom near the teacher’s desk. Six iPads were placed upon the back three student desks. The classroom has a closed, dedicated AHU providing mixed fresh and recirculated air through three variable air volume (VAV) boxes. Four linear diffusers next to the windows and three square diffusers in the center of the classroom supplied air (Figure 1b), with two return grilles at the northwest and southwest corners. A minimum efficiency report value (MERV) 10 filter, common to public spaces, including schools, was installed in the AHU. The MERV 10 filter has an average particle size efficiency of 50% - 64.9% for particles sized 1.0 - 3.0 micrometers and 85% or greater for particles 3.0 - 10.0 micrometers.^47^ During each experimental condition, the classroom was maintained between 20 and 22 ℃ and 20-50% RH.

**Figure 1.**
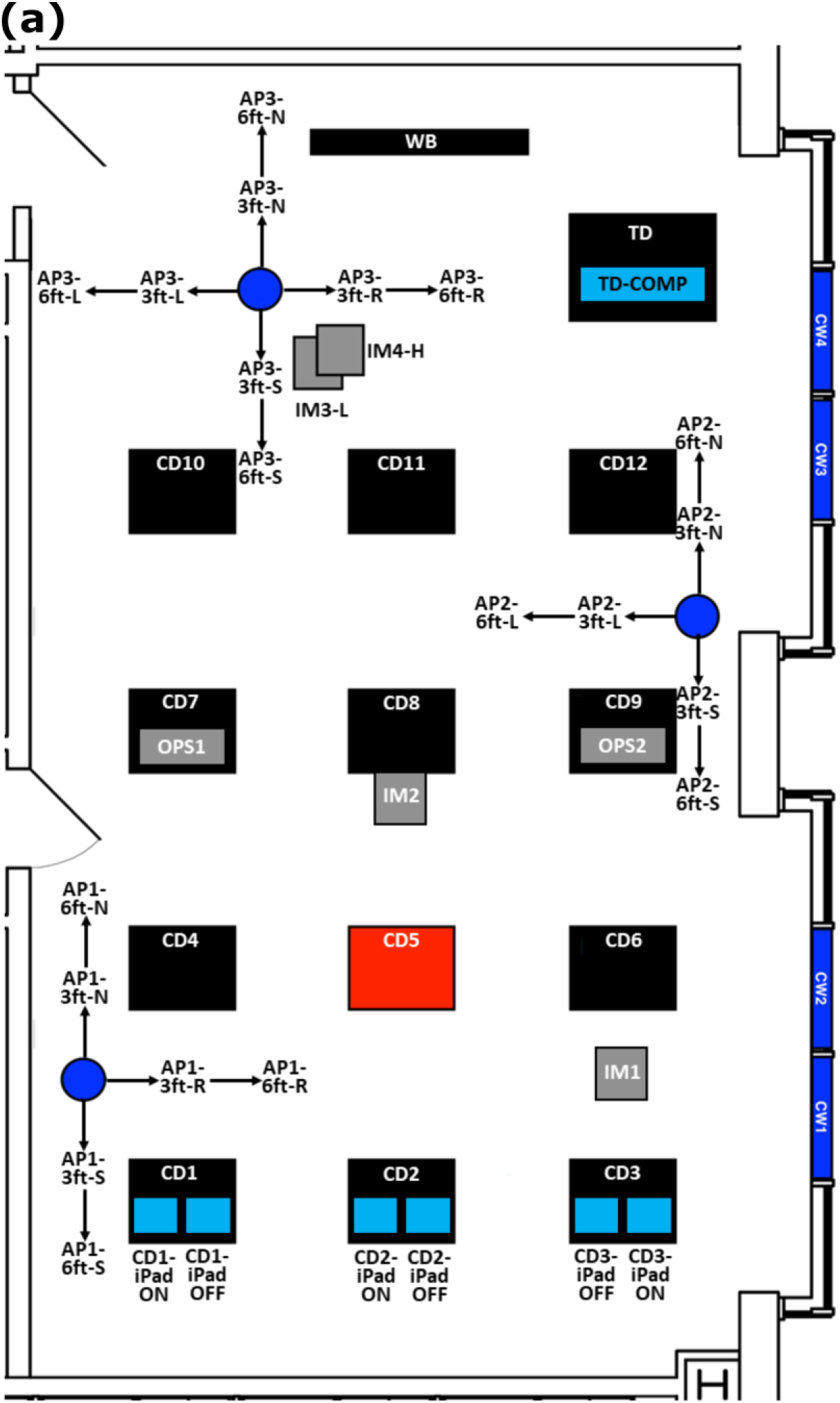
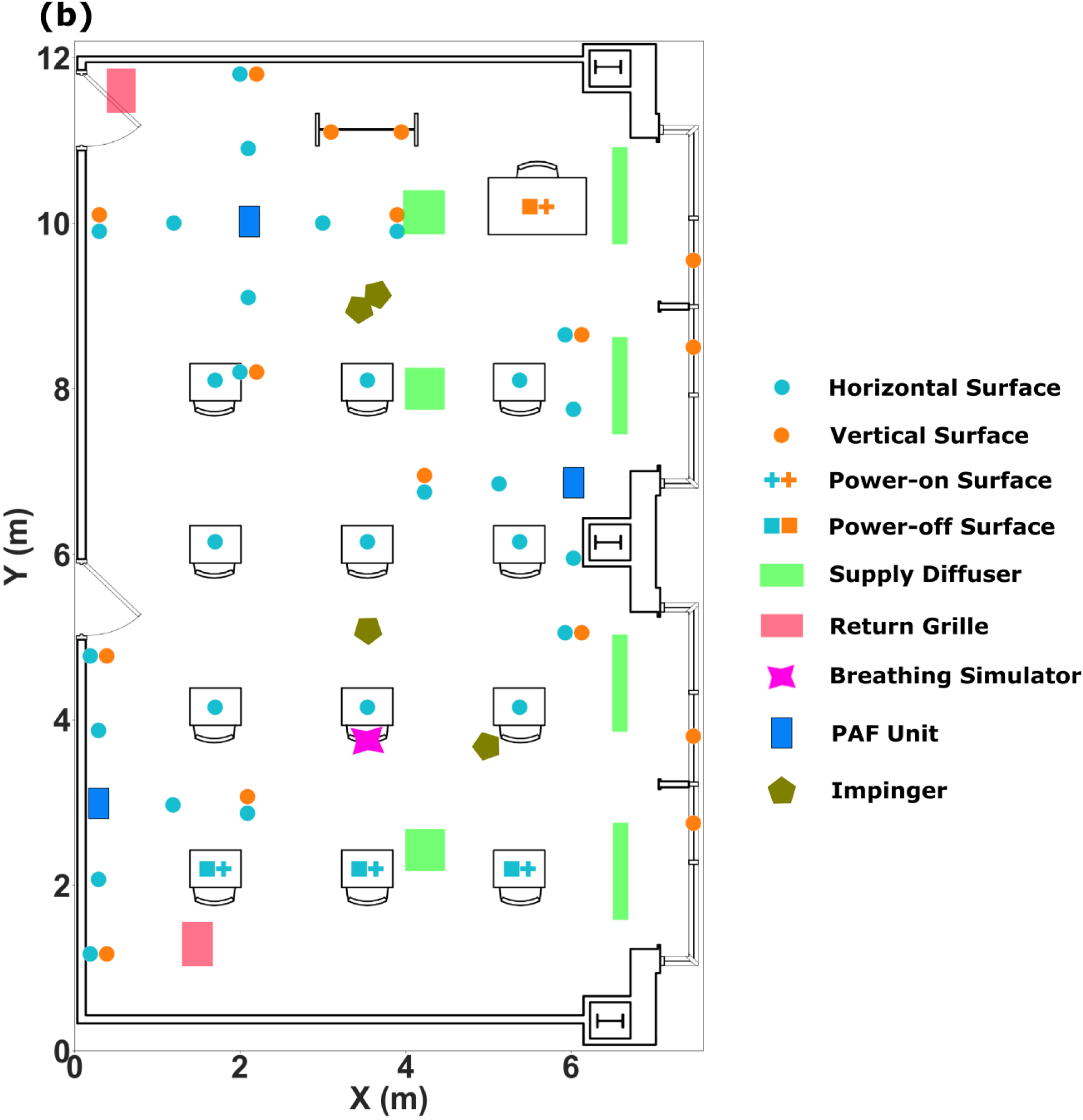
A schematic diagram of the classroom environment utilized for measurements **(a)** and locations of each sampling device/substrate **(b)**. (CD-classroom desk; SIM-breathing simulator; OPS-optical particle sizer; IM-impinger; PAF-portable air filtration unit; CW-classroom window)

The ventilation rate to the classroom by the AHU was designed and set based on ASHRAE Standard 62.1^48^ and intended to mimic common U.S. classrooms. Prior to experimental measurements, we performed a CO_2_ decay test using an ASTM international standard for determining air change in a single zone^49^ in order to quantify the total air supplied to the room (ventilation + infiltration/exfiltration). Briefly, we placed two tanks of CO_2_ in the classroom each injecting 68 m^3^/h (40 CFM) of CO_2_. We used two large fans to help mix the classroom air. The uniformity of the CO_2_ distribution was confirmed in previous tests. A TSI Q-Trak Probe 982 (TSI Inc.; Shoreview, MN) was used in this study to monitor the CO_2_ concentration change in the classroom, with the sensor placed in the middle of the classroom and set to a sampling interval of one second. Injection continued until the concentration of the CO_2_ reached 3000 ppm and stablized. After the CO_2_ was shut off, we monitored the CO_2_ concentration until near-background levels (350-600 ppm) were acheived. Using these CO_2_ decay data, the air change rate *A* could be determined using via the equation:^49^

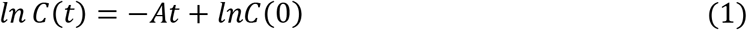

where *C*(*t*) is the CO_2_ concentration at time *t*, and *C*(0) is the CO_2_ initial concentration when the decay started. We completed the CO_2_ decay test in triplicate, with the mean total air change rate of the room observed to be 3.33 ± 0.05 h^-1^ (766 ± 10 m^3^/h).

### 2.2. PAF Unit Description

Three PAF units were also placed in the classroom (Figure 1), spread nearly equadistant in the space to mimic placement in a typical classroom while still allowing for our measurements to be made properly. Each PAF unit was an Intellipure Compact (Intellipure, Inc.; Pulaski, New York), which contains a six-stage prefilter for larger particles, and VOCs, followed by an electrostatic precipitator (ESP) and, subsequently, a dielectret main filter for micrometer and submicrometer particle collection. While we did not perform tests to directly evaluate particle collection efficiency within these units,^38^ we remark that the multiplexing of different technologies in series in PAF units is commonly done in an effort to maximize collection efficiency while simultaneously minimizing pressure drop.^50^ In series, application of multiple control technologies reduces the influence of leaks, which have been detected in prior studies^51^ of PAF units incorporating single technologies. In particular, the combination of unipolar ionization, or an ESP, with a filter has been shown to yield enhanced collection efficiency in HVAC systems employing lower MERV rated filters.^50^ Within the ESP, there is a wire-based unipolar ionizer generating nominally 1,750,000 positive ions/cm^-3^ just prior to the dielectric main filter, with minimal ozone generation (<0.001 ppm) and close-to-zero (undetectable) ion escape outside the unit. We have included further information on anticipated diffusion and field charging levels in the ESP in the supporting information (Sec. 2) along with information on the air flow speeds from the unit when run with and without filters installed (Table S1 and Figure S1 in the supporting information). At a particle size of 0.3 micrometers and using the testing standards EN1822-1:2009^52^ and EN1822- 5:2009^53^, the particle removal efficiency was 99.83% for the filter within these units and higher for other sizes^52, 53^ (manufacturer documentation). When the whole unit was tested as an integrated filter + ESP according to these testing standards, the particle removal efficiency was 99.997% at 0.3 micrometers (manufacturer documentation). When run on the highest fan setting, each unit had a third-party tested CADR of approximately 246.9 m^3^ hr^-1^. The three units hence led to an augmentation of ∼3.23 ACH within the classroom, yielding a total effective ACH of 6.56 assuming addivitiy with the HVAC system.

**Table 1.**
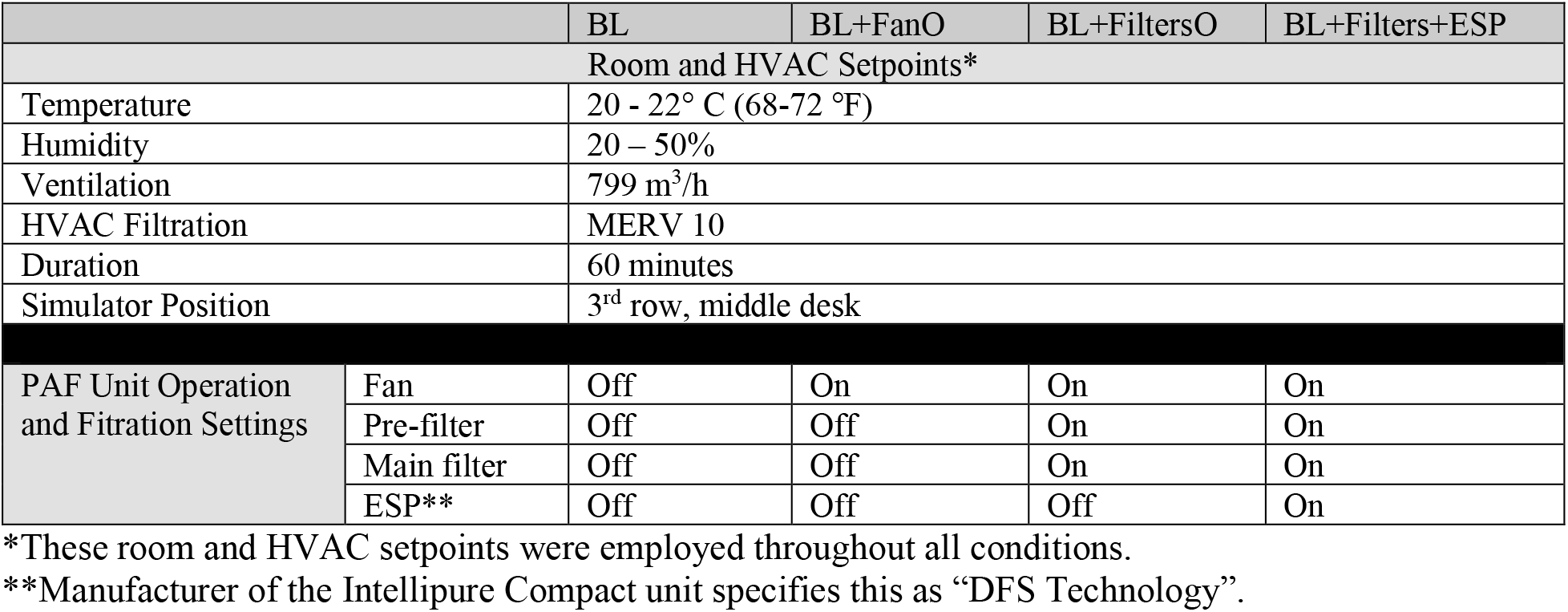
Summary of the experimental conditions employed in trials

### 2.3. Measurement Methodology

A breathing simulator (Figure 2a) was placed in the classroom (middle desk in the 3^rd^ row in Figure 1) to mimic the breathing of a 1.6 m height, 49 kg female^54^, best approximating a high school student. The simulator was comprised of an anatomically correct manikin (Mayo Clinic Respiratory Care, Laerdal) whose respiratory anatomy was connected to a Blaustein Atomizing Modules (BLAM) nebulizer. The breathing simulator development, setup, and validation, as well as the particle deposition assessment protocol, has been outlined within Eilts et al.^20^ Briefly, we used breathing waveform equations from Gupta et al.^54^ to ensure proper inhalation and exhalation from the simulator. The inhalation and exhalation waveforms of the simulator were 0.38sin(1.67*t*) and 0.32sin(1.39*t*) standard liter per minute (slm), where *t* is the time in seconds, *α* is the amplitude in L/sec, and *β* is the frequency in sec^-1^.^54^ The nebulizer generated aerosol using a solution of 20% glycerol, 1.5% uranine (fluorescein sodium salt), and 78.5% distilled water under a constant backing pressure of 2.07 bar that was provided by an air compressor. The mean volumetric particle size was approximately 1.8-2.0 μm by mass after evaporation. During inhalation, particles produced by the nebulizer were passed to a filter and not expelled into the room, while during exhalation the rate of particle dispersion followed the exhalation waveform.

**Figure 2.**
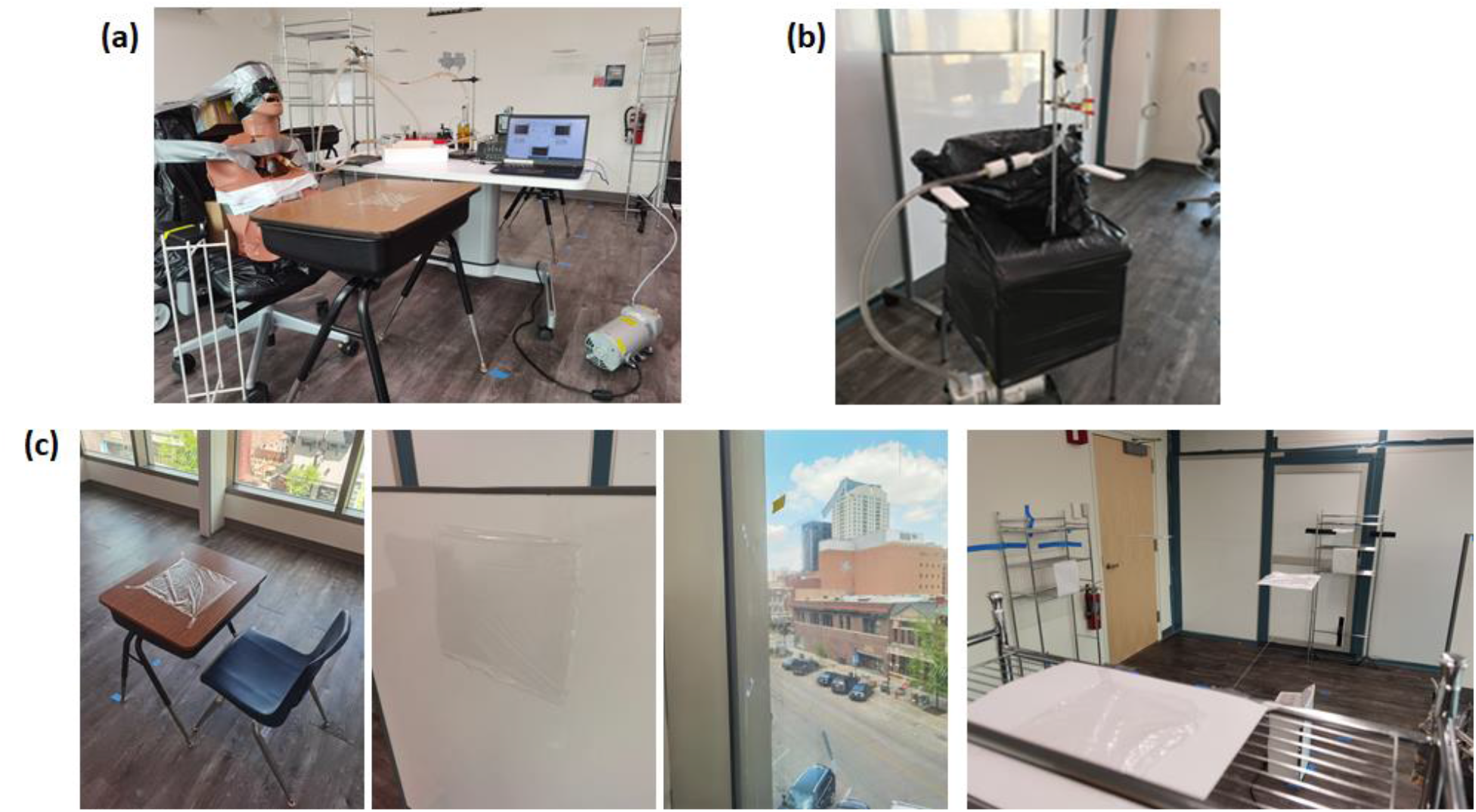
Photograph of the beathing simulator (a), the impinger samplers (b), and saran used in deposition assessment (c).

To examine particle dispersion, we used a combination of techniques. Two opticle particle spectrometers (OPS; TSI 3330; TSI Inc., Shoreview, MN) measured the particle concentration and size distributions at two different locations (Figure 1) in the 0.3 – 10 μm size range. Both OPS locations were one row in front of the simulator, but one OPS was next to a PAF unit in the middle right of the classroom (AP2) on CD9 (child desk) while the other OPS was distal to all PAF units, on CD7. Four impinger sampling systems were also utilized to measure the fluorescein-tagged particle mass concentration at 1) different heights around the PAF unit most distal to the simulator (IM3-L and IM4-H) and 2) near the simulator (IM1 and IM2). Each sampling system consisted of a small glass impinger (AGI-30; Ace Glass, Vineland, NJ) and a HEPA capsule connected via Tygon tubing to a vaccuum pump (McMaster Oil-Free Electric Vacuum Pump; Santa Fe Springs, CA) (Figure 2b). Impingers were operated at 12.5 liters per minute sampling rate, and were filled with 25 ml NaOH solution (0.001 M, mixed with 4 g sucrose to prevent freezing). Three of the four impingers (IM1, IM2, and IM4-H) were placed 1.52 m above the floor to sample the air from the breathing zone expected when an average height person is standing, while IM3-L, colocated with IM4-H, was placed 0.76 m above the floor. This lower height allowed for an assessment of the impact of the PAF unit vertically. The vacuum pumps used for operation were placed underneath tall chairs covered with 55-gallon plastic bags to limit any impact of thermal plumes generated by the running of the pumps.

Saran wrap and scotch tape were used as substrates to collect and measure fluorescein-tagged particle deposition onto various vertical and horizontal surfaces in the classroom. Saran wrap was placed on: 1) the students’ (×9) and teacher’s (×1) desks; 2) the white board (×2) in the front of the classroom; 3) the windows (×4); and 4) suspended paper platforms strung between and placed upon wire shelving units at 0.91- and 1.83-m distances from each PAF unit, with these platforms 1.5 m above the floor in the breathing zone expected when an average height person is standing (×30). Images of selected substrates are shown in Figure 2c. The saran wrap pieces were smaller (15.24 cm × 15.24 cm) on surfaces within 2 m of the breathing simulator and around all PAF units on the suspended paper platforms and wire shelving units, with larger saran wrap pieces (30.48 cm × 30.48 cm) for surface locations greater than 2 m from the breathing simulator. Examining whether the electrostatic impact of electronic devices increases particle deposition to these items was also of interest, as during preliminary investigations of our prior study, we observed preferential deposition of microparticles on ungrounded, statically charged surfaces.^20^ Therefore, deposition to the screens of electronics in the classroom was examined via collection on screen protectors placed on the computer monitors (×2) on the teacher’s desk and iPads (×6) on the last row of student desks, with two iPads on each of these desks. One iPad was powered on and the other powered off on each desk. Finally, we placed pieces of scotch tape (8.9 cm × 1.9 cm) on the supply and return grilles to measure deposition to these surfaces. Unlike the other deposition surfaces, which can be described as passive in their mechanism of particle collection, the deposition to the supply diffusers and return grilles was active, as the air was either being pushed away or pulled into, respectively, these surfaces, enabling collection by interception and impaction.

### 2.4. Experimental Procedures

To examine the impact of PAF units, we implemented four experimental conditions (Table 1). The four conditions were:

(1) *BL*: Baseline HVAC Only
(2) *BL+FanO*: Baseline HVAC+Fan Only PAF unit operation [filters removed and ESP powered off]
(3) *BL+FiltersO*: Baseline HVAC+Filters Only PAF unit operation [ESP powered off]
(4) *BL+Filters+ESP*: Baseline HVAC+Filters and ESP PAF unit operation.

Each trial was initiated with a thorough cleaning of the classroom to reduce background levels of fluorescein-tagged particles in the air and on surfaces. All surfaces were cleaned using disinfectant wipes, and the classroom air was scrubbed for more than 24 hours by the room’s HVAC system and a standalone commercial PAF unit with a CADR of 1614 m^3^/h (950 CFM). We then completed an 18-hour control test where we placed a subset of saran wrap substrate throughout the classroom on desks and windows as well as scotch tape on all supply diffusers and return grilles. After this 18-hour period, the substrates were collected and processed to determine backgroud levels (processed identically to the procedures described below). The control conditions showed background deposited fluorescein masses 2-4 orders of maginitude below that measured during conditions where the breathing simulator was present and operating, and was corrected for in data processing.

Following control measurements, the breathing simulator, PAF units, OPS units, impingers, and surface substrates were then placed into the classroom according to Figure 1, with their operation and filtration status following the specifications in Table 1. For the baseline condition without the PAF units operating, these units were still placed in the same place with the inlets and outlets sealed. During conditions with the PAF units running, we operated these units at their highest setting, *i.e.*, with a CADR of ∼246.9 m^3^ hr^-1^ per unit. Between conditions, we cleaned the units carefully with disinfectant wipes to minimize any background flourescein-tagged particles the units themselves may introduce. All deposition experimental conditions were completed without Researchers within the classroom aside from brief entry into the classroom at the start and at the conclusion of each condition. At the start of each experimental condition, we implemented a 15-minute period where only the two OPS units within the classroom were running. This collection period allowed for the collection of particle background data for each experimental condition. After this particle background data collection period, we then completed a 60-minute injection with the breathing simulator, impinger sampling systems, and OPS units all powered on simultaneously. For the experimental conditions with the PAF units operational, these units were also powered on at the same time as the preceding components. At exactly 60 minutes, we powered off the breathing simulator and impinger sampling systems but left the OPS units to run for another 15-minute period for the BL and BL+Filters+ESP conditions. These final 15-minute periods allowed for collection of particle decay data, akin to CADR testing but not in a sealed environment. Even after the 15-minute decay period, the HVAC system and the PAF units (if used during the condition) remained on and at the same settings. They remained in this setup until all processing (outlined below) was completed.

The measurement procedure was immediately followed by the processing of all surface substrates and the solution within the impingers. We removed each piece of saran wrap and scotch tape one-by-one from the classroom, with each piece of substrate placed in an individual sterile petri dish for processing. The fluorescein on each substrate was recovered by scraping the surface of each piece of substrate exposed during the condition with a sterile cell scraper or sterile cotton applicator (for screen protectors on electronic devices only), with the buffer solution used during these extraction procedures being 3 mL of 0.001M NaOH per piece of substrate. Prior to processing each piece of substrate, we cleaned the processing table with a disinfectant wipe, and we used a new sterile cell scraper or cotton applicator as well as a new pair of gloves to process each substrate. This minimized potential cross-contamination. The collected solution was then pipetted into a new 12 x 75 mm disposable glass culture tube for each piece of substract, and its fluorescein concentration was determined using a Trilogy Laboratory Fluorometer previously calibrated using a set of standard fluorescein solutions of known concentration. Each impinger was reweighed after being removed from the classroom, with the pre- to post-test impinger weight difference yielding the loss of sampling liquid due to evaporation. The volume of the liquid left in each impinger was also recorded, and the fluorescein concentration in the remaining liquid was measured by pipetting 1 mL of the remaining solution from the bottom of each impinger into new disposable glass culture tubes and placing into the Fluorometer. Following processing, the HVAC system and the PAF units were powered down or off, respectively.

Along with the combined particle concentration and deposition measurements, we conducted four additional experimental conditions to determine the aerosol concentration distribution within the classroom, again using the settings specified in Table 1. For these measurements, we placed one OPS unit on the middle student desk one row in front of the breathing simulator (CD8) as a reference, with the other OPS unit placed upon a cart 81 cm in height and moved every 2 minutes to the 34 locations indicated by the blue squares in Figure S4 of the supporting information. The cart had a wire frame that minimally impeded air flow. For each measurement, we first ran the breathing simulator for 60 minutes to inject the classroom with enough aerosol to reach a steady state. After 60 minutes, two Researchers entered the classroom. One Researcher was situated at the side of the classroom with a study laptop and remained in this location throughout the duration of the experiment. The other Researcher guided the OPS unit situated on the cart already positioned at location “1-CD3-O” to each of the 34 sampling locations denoted by precisely measured pieces of tape on the floor. With the breathing simulator still running, we sampled at each location for a total of two minutes, with the OPS units sampling at 10-second intervals. Each location’s ‘start’ and ‘stop’ times directly from the OPS unit being moved around on the cart were stated by the Researcher pushing this cart for documentation into a spreadsheet by the Researcher holding the study laptop at a fixed location in the classroom. The data were cleaned to remove the outliers outside the 95% confidence intervals. The concentration and size distribution of the aerosol particles at each location facilitated analyses of the spatial effect of the PAF units on particle concentrations throughout the classroom under each experimental condition. More detailed procedures can be found in the Section 3 of the supporting information.

### 2.5. Data Processing

The raw fluorescein mass concentration recovered from the substrates and impingers was recorded in mg/m^3^. Extraction efficiency experiments for each type of substrate were performed in tripicate using 0.1 mL of 100 mg/m^3^ fluorescein solution in a room separate from the classroom using a different AHU. After a drying period of 24 hours, we processed these pieces of substrate identically to that outlined above. We then used the extraction effiency as a correction factor for each type of substrate; the fluorescein mass concentration *c_d,i_* in the recovery solution after processing each piece of substrate was determined by dividing the raw concentration by its extraction efficiency. The effective deposition fluxes *J*" for each substrate were then calculated as:

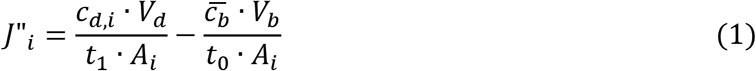

where *V*_*d*_ is the volume of the NaOH solution used for recovery, 3 × 10^−6^ *m*^3^; *t*_1_ is the duration of the injection, 3,600s; *A*_*i*_ is the area of each substrate, 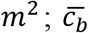 is the averaged fluorescein concentration of the background substrate recovery solution, *mg*/*m*^3^; *V*_*b*_ is the volume of the NaOH solution used for background recovery, 3 × 10^−6^ *m*^3^; *t*_1_ is the duration of the background test, 64,800s. Then, for each substrate, the deposition velocity ℎ_*i*_ at location *i* can be determined using the following equation,^25^

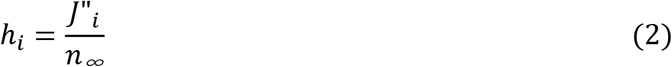

where *n_∞_* is the aerosol mass concentration of fluorescein particles at the height of the simulator, determined as the average from the three impingers situated at the height of the breathing zone expected when an average height person is standing (IM1, IM2, and IM4-H).

## 3. Results

As prior studies have examined particle concentration decay in classroom settings via implementation of PAF units,^40, 42^ we elect to focus first on examining PAF unit influence on surface deposition. We separate passive horizontal upward-facing surfaces, passive vertical surfaces, active horizontal downward-facing surfaces, and electronic surfaces in this comparison, as the first three were anticipated to have different rates of deposition based on the possible deposition mechanisms which can drive particles to these surfaces.^22, 25^ Concentration changes and clearance data are subsequently presented to provide a holistic view of PAF influences.

### 3.1. Deposition to Passive Horizontal Upward-Facing Surfaces

Deposition velocities to all passive horizontal upward-facing surfaces (including desks and paper platforms strung between and on wire shelving units) at different locations throughout the classroom during each experimental condition are plotted in Figure 3a. Deposition velocities ranged between 10^-4^ and 10^-1^ cm s^-1^. For most passive horizontal upward-facing surfaces, the BL and BL+FanO conditions had comparatively higher deposition velocities than the BL+FiltersO and BL+Filters+ESP conditions, with consistent notable spikes at CD5-SIM. CD5-SIM represents deposition to the student desk immediately in front of the breathing simulator, so deposition velocity was typically higher than at other locations, as expected. While spikes were observed at the AP1-3ft-R and AP3-6ft-N locations, this appeared only during the BL+Filters+ESP condition and may have been due to changes in localized airflow, an effect we discuss subsequently. Figure 3b plots deposition velocity for all passive horizontal upward-facing surfaces by distance from the breathing simulator. As expected, and consistent with our prior work examining deposition in an office environment,^20^ the deposition velocities were highest within 2 m from the breathing simulator and stabilized between 10^-3^-10^-2^ cm/s beyond 2 m. This suggests that the indirect transmission route via deposited aerosol particles, if significant, would be most likely to occur through contact with surfaces that an infected individual remained close to (within 2 m of) for an extended period of time. Further, the slight downward trend as distance from the breathing simulator increases suggests a change in mass transfer by distance and that perhaps the well mixed assumption^55^ needs to be employed with caution when understanding the transport of particles within an indoor space.

**Figure 3.**
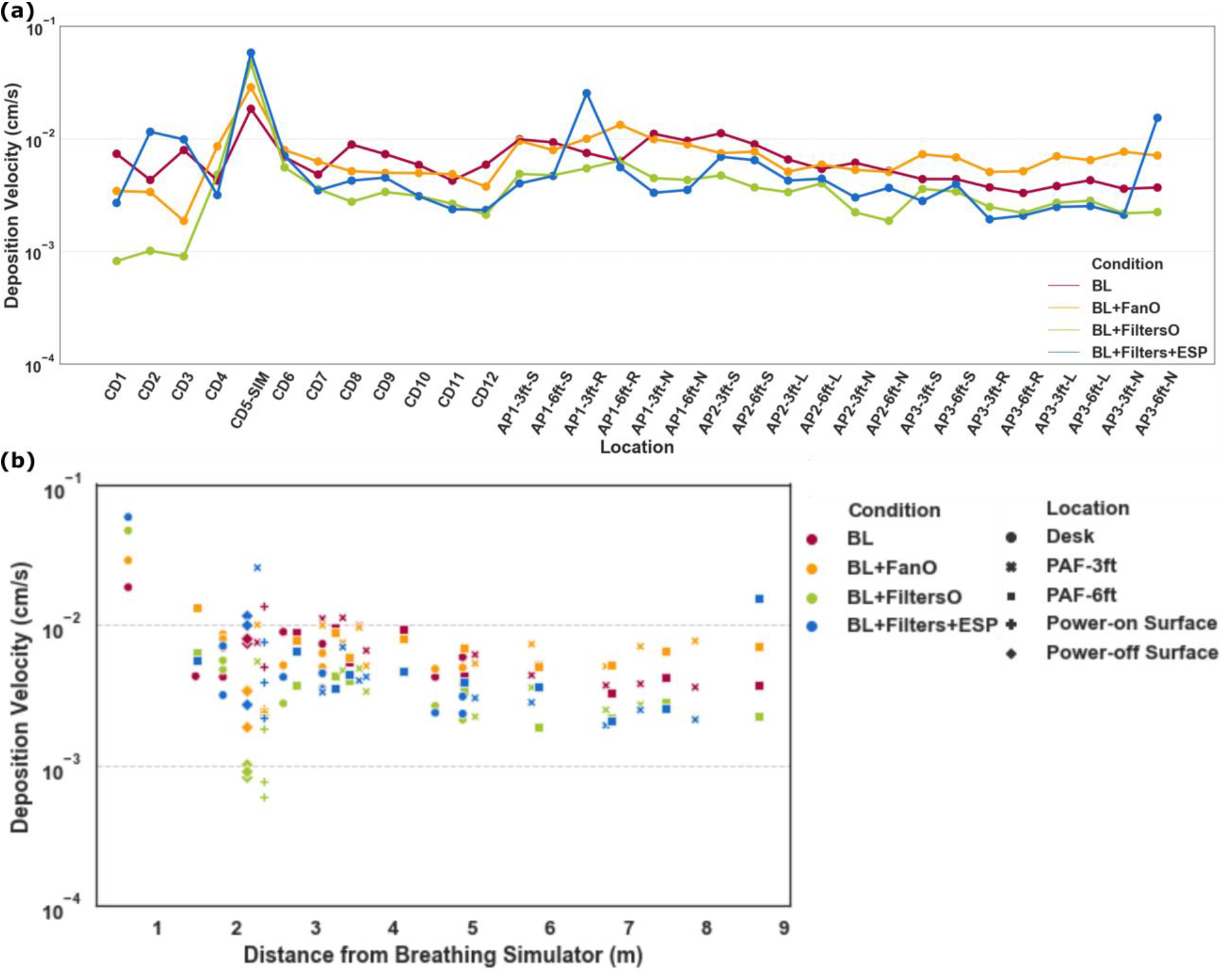
Deposition velocities under different conditions plotted by labeled passive horizontal upward-facing surfaces **(a)** and by distance from the simulator **(b)**.

### 3.2. Deposition to Passive Vertical Surfaces

Deposition velocities for all passive vertical surfaces are plotted in Figure 4a by distance from the breathing simulator. Similar to the passive horizontal upward-facing surface deposition data, the BL and BL+FanO conditions typically had higher deposition velocities than the BL+FiltersO and BL+Filters+ESP conditions for passive vertical surfaces. However, relative to passive horizonal upward-facing surfaces, the deposition to passive vertical surfaces was appreciably slower at 10^-5^-10^-2^ cm s^-1^ and demonstrated greater scatter. Figure 4b compares deposition velocity for the passive vertical surfaces (1.23 m above the floor) situated 1.83 m from each PAF unit to the passive horizontal upward-facing surfaces (1.52 m above the floor) at the same distance from each unit at the same location, with significantly lower (p ≤ 0.01, two-sided t-test with unequal variances) deposition velocity for all passive vertical surfaces.

**Figure 4.**
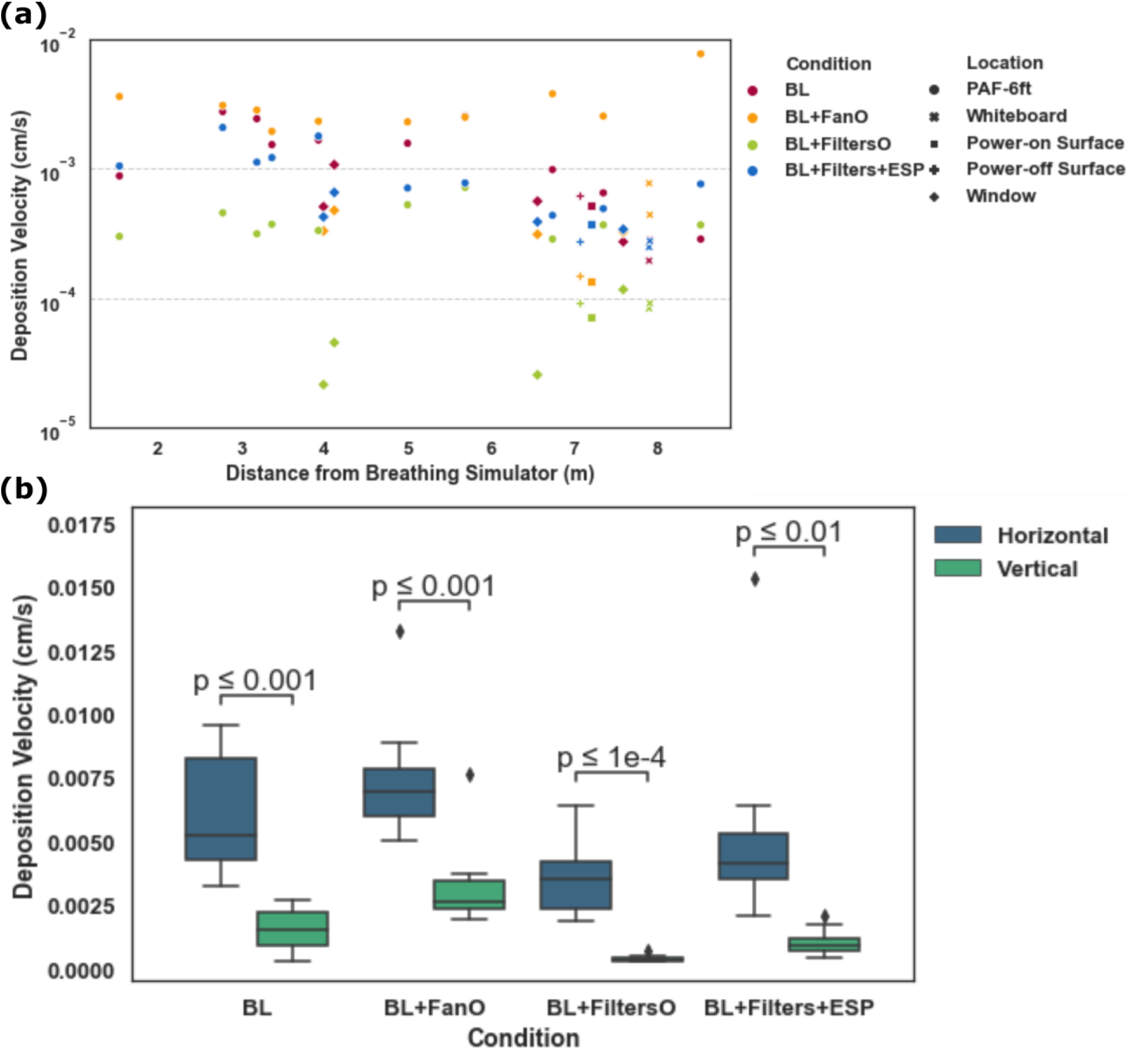
Vertical surface deposition velocities as a function of distance from the breathing simulator **(a)**. Comparison of the deposition velocities for passive horizontal upward-facing surfaces and passive vertical surfaces for the four examined measurement conditions **(b)**.

These observations for passive horizontal upward-facing surfaces and passive vertical surfaces agree largely with Lai and Nazaroff’s framework^25^ for modeling particle deposition onto differently oriented surfaces. For microparticles, the influence of gravity cannot be neglected for deposition onto upward-facing surfaces. For characteristic velocities of indoor air, their model indicates that the deposition velocities of particles 0.1-10µm in size onto passive vertical surfaces spans the 10^-6^-10^-4^ cm s^-1^ range, while the deposition velocities onto passive horizontal upward-facing surfaces varied between 10^-4^ and 10^-1^ cm s^-1^. Although deposition velocities onto passive horizontal upward-facing surfaces in our study agreed well with their model, we observed higher deposition velocities onto the passive vertical surfaces than they referenced. As pointed out by our previous study^20^ the reason for this difference can be attributed to the effect of the ventilation system on the room’s flow field, surface roughness, and the use of the reference concentration for a point source, as well as a potential limitation of the measurement device.

### 3.3. Deposition to Active Horizontal Downward-Facing Surfaces

Figure 5 displays deposition velocity to active horizontal downward-facing surfaces, which are diffusers and return grilles, by distance from the breathing simulator. As expected, deposition was slightly higher for the return grilles relative to the diffuser given that these surfaces were pulling air from the classroom versus supplying the classroom with air, respectively. These data generally suggested that for the BL and BL+FanO conditions: (1) more particles needed clearance by the HVAC system due to particles not being filtered and then being recirculated back into the classroom by the PAF units (either because they were off [BL condition] or because they had no filters and no ESP [BL+FanO condition]); and (2) more particles were being recirculated back into the classroom through the HVAC system because, again, more particles were entering the HVAC system in the first place. Additionally, and consistent with our prior measurements,^20^ deposition on these surfaces show little relationship with distance from the breathing simulator, with much higher deposition velocities observed even 8 m away, than were observed on passive vertical surfaces less than 2 m from the simulator.

**Figure 5.**
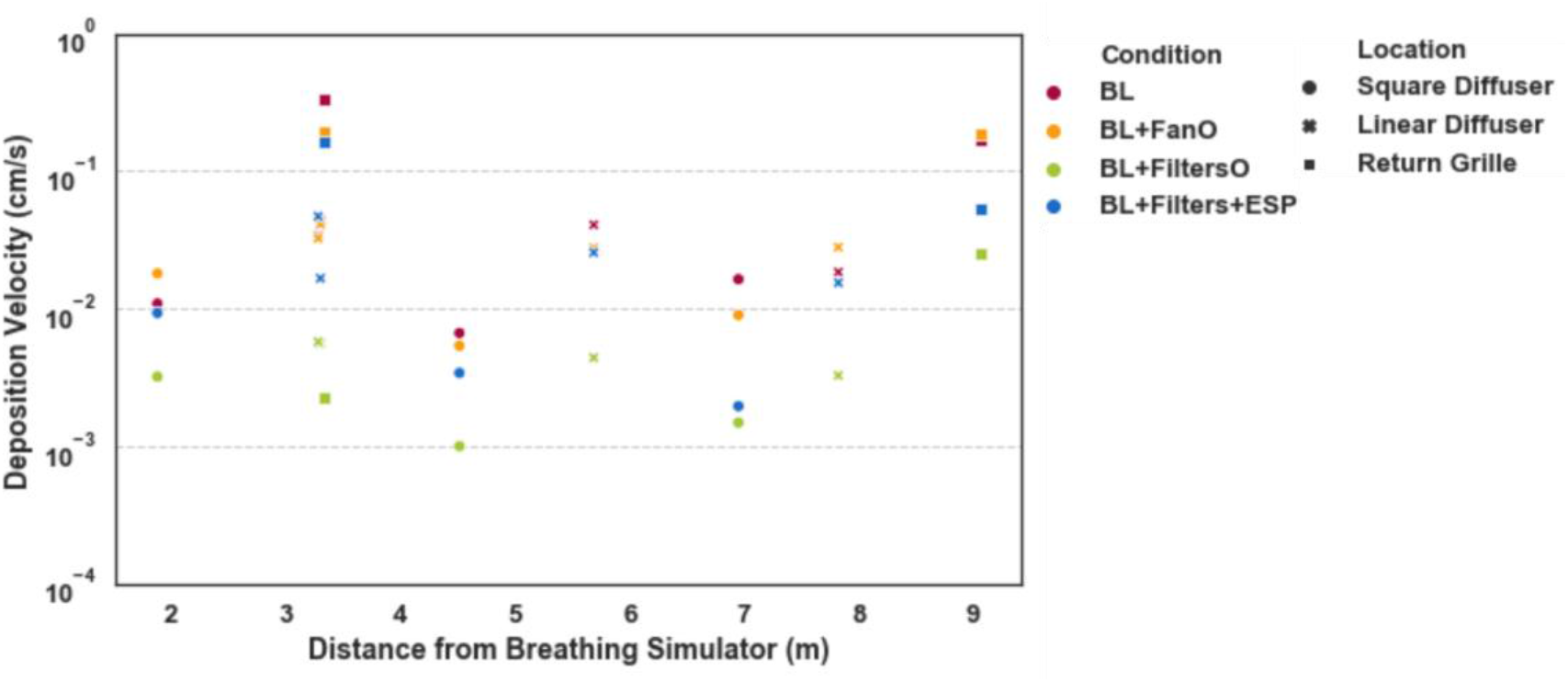
Deposition velocities for active horizontal downward-facing surfaces by distance from the simulator.

### 3.4. Deposition to Electronic Surfaces

Electrostatic forces have been found to significantly increase particle deposition in the indoor environment, as a strong electric field can be present close to electronic screens, with particles also able to gain a charge via collision with air ions.^56^ Furthermore, in the present experiments, we utilized nebulized particles without neutralization (prohibited by the high flow rates of the breathing simulator relative to the flow rates of the bipolar ionization sources we had at our disposal). Particles therefore were likely charged considerably from the aerosolization process itself. We thus compared particle deposition to iPads and computer screens powered on and powered off in colocated areas of the classroom, to determine the extent to which electrostatics had an influence on experimental results, which may influence particle deposition in a classroom setting. A summary comparison is shown in Figure 6, revealing there were no significant differences in deposition velocity between electronic devices powered on or off. We attribute this finding to little change in the iPad and computer monitor surface charge distributions and electric fields when powered on compared to powered off scenarios, particularly compared to the effects of fluid flow and gravity on particle motion in the classroom environment. This finding can perhaps be generalized to electronics in a classroom setting; powered systems will not necessarily lead to different levels of particle deposition than non-powered systems. We believe more research will be needed to understand the influence electrostatics may play in indoor air deposition, including not only electronics, but also static charge levels on different building materials, and the variation in these charge levels with temperature and relative humidity (*i.e.*, with season and time of day).

**Figure 6.**
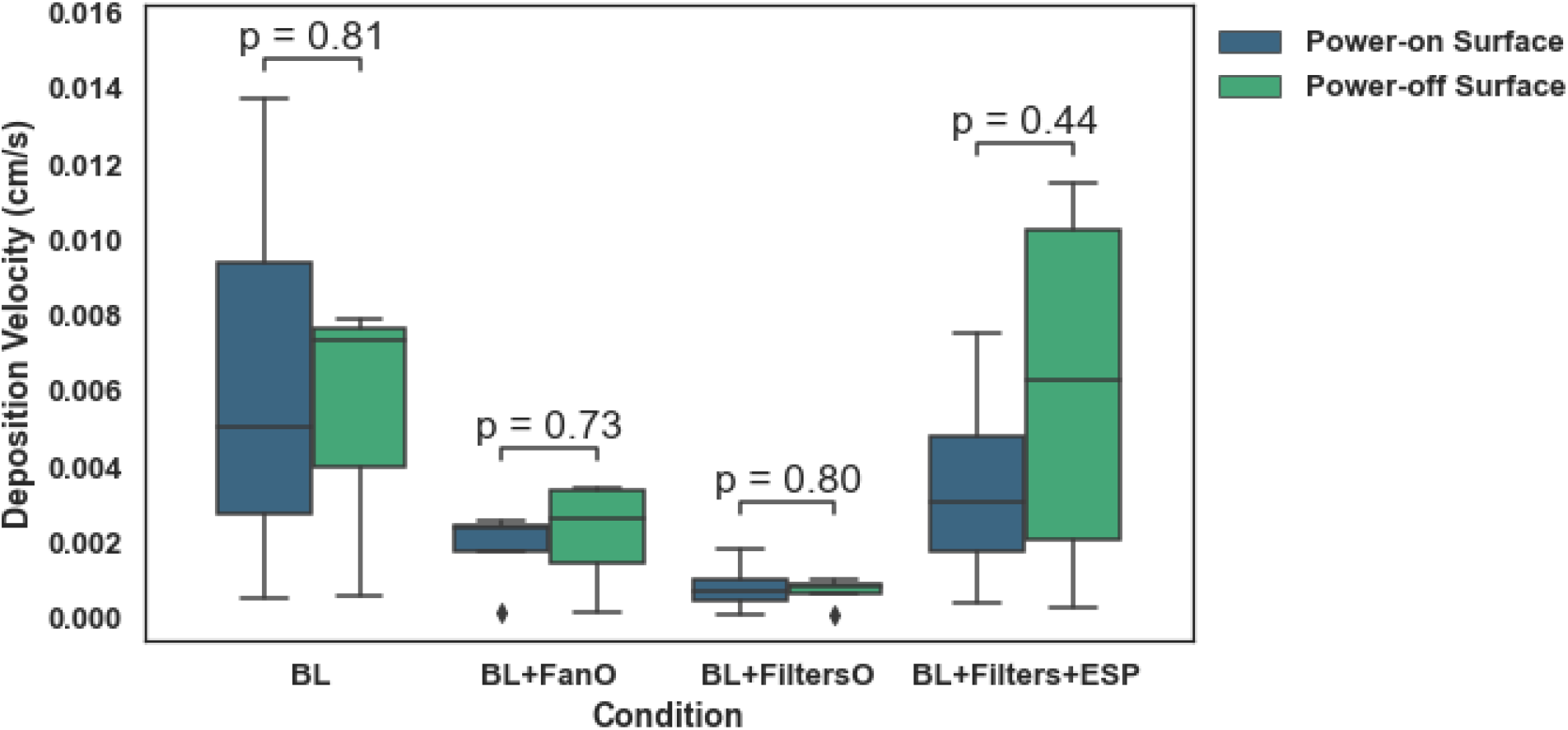
Comparison of deposition to electronic surfaces powered on and off by condition.

### 3.5. Aerosol Concentration Distribution and Decay

While deposition velocity is important when discussing the potential for indirect transmission, the total aerosol particle concentration can serve as a proxy for the risk of direct inhalation of potentially infectious airborne particles. Total particle concentrations in the classroom for the different conditions are plotted in Figure 7a in the 0.3 – 10 μm size range. Average concentrations at different classroom locations ranged from ∼100 to 1100 #/cm^3^. As reviewed in our methodology section, these experimental conditions were completed separately from the deposition conditions, but with the room, HVAC, and PAF unit setup being identical. As expected, higher concentrations were observed at the locations close to the breathing simulator, such as CD5-SIM, CD6, and AP2-6ft-N. However, this trend was not consistent between different conditions. For example, the BL case had a higher concentration at AP-2-6ft-S, which was to the back left of the simulator, while the BL+FanO condition had higher concentrations at CD5 and CD6, which was directly in front of and to the right of the simulator. This suggests that the concentration of aerosol particles throughout the classroom was influenced by changes in airflow due to PAF unit operation. We also remark that these concentrations are not background corrected and are hence a combination of breathing simulator generated particles and background aerosol. Encouragingly, the BL+FiltersO and BL+Filters+ESP conditions had aerosol particle concentrations that trended lower throughout most locations of the classroom relative to the BL and BL+FanO conditions, particularly at distances beyond 2 m from the simulator. The averaged aerosol particle concentration values demonstrated lower peaks and narrower ranges during the BL+FiltersO and BL+Filters+ESP conditions, even within the vicinity of the breathing simulator. Overall, the particle concentration with the PAF units operating with both filters and the ESP reduced peak concentrations by a factor of around 2.5 compared to the highest concentrations observed, which were with the BL+FanO condition.

**Figure 7.**
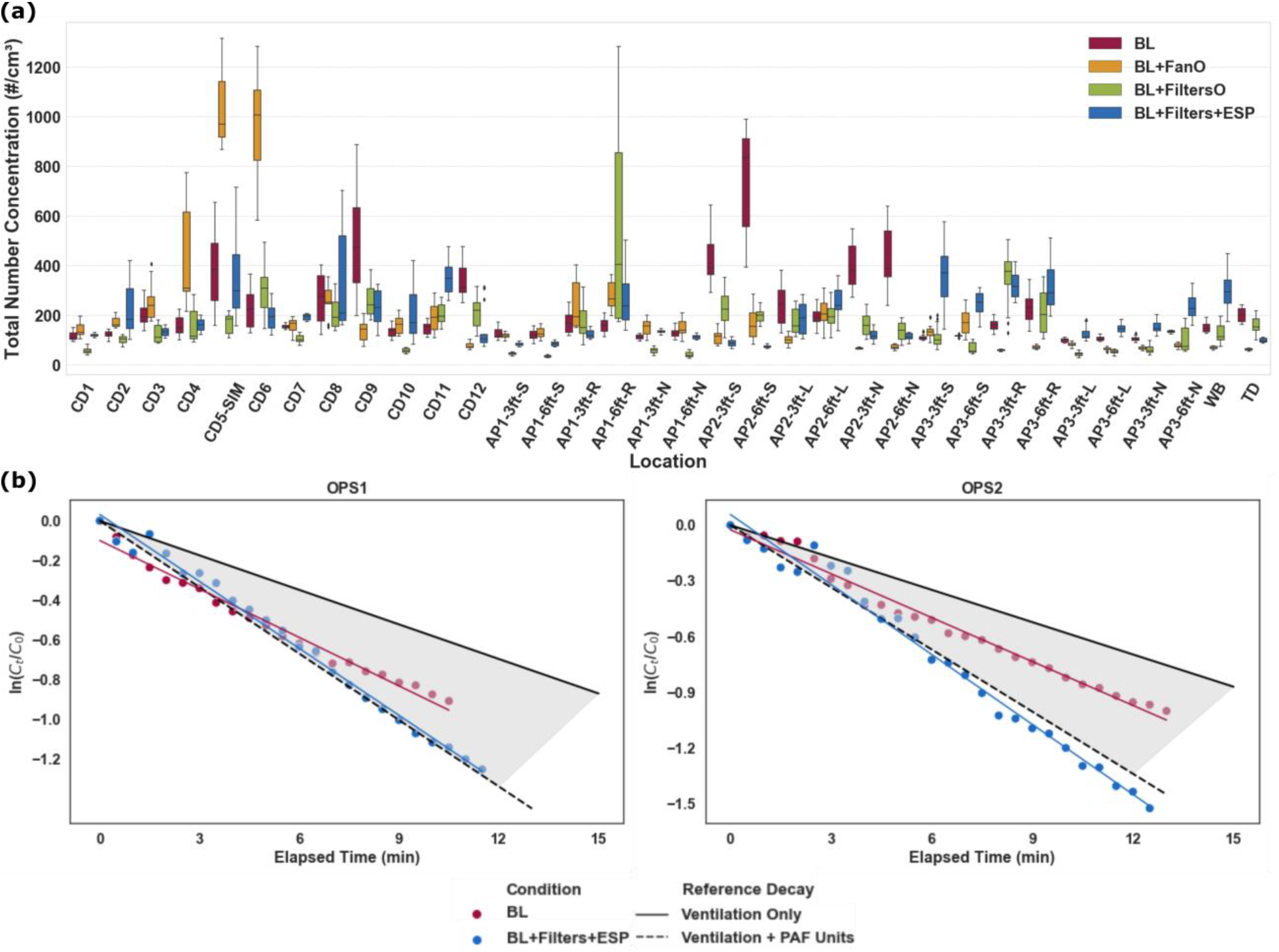
Total particle number concentrations at different locations of the classroom by condition **(a)** and the normalized particle concentration decay for the four measurement conditions, in comparison to predictions based upon air change rate **(b)**.

As described in the “Methods” section, the BL and BL+Filters+ESP conditions afforded the opportunity to examine particle concentration decay and compare them to expectations based upon the air change rate from the ventilation system. The total particle decay data, presented as the natural log of the particle concentration divided by the concentration at the time the breathing simulator was shut off in the 0.3 – 10 μm particle size range, are plotted in Figure 7b. Also plotted is the expected decay curve neglecting deposition but considering the average measured air change rate for the AHU, and the augmentation brought about by the PAF units. Immediately evident is that for the data from both OPS systems, results are in good agreement with theoretical expectations for the BL+Filters+ESP condition, i.e. the PAF units led to the anticipated level of particle clearance based upon their CADR values. This is consistent with recent studies of PAF unit influences on aerosol concentration decay, both experimental^40^ and numerical^57^.

## 4. Discussion

### 4.1. Effects of PAF Units on Deposition

From measurements, it is clear that passive horizontal upward-facing surfaces have much higher deposition velocities than vertical passive surfaces (by more than 2 orders of magnitude in some circumstances). Therefore, in terms of mitigating indirect transmission, it is these surfaces that are of concern. We do find that PAF operation with control technology implemented (*i.e.*, the BL+FiltersO and BL+Filters+ESP conditions) led to reduced deposition velocities on passive horizontal upward-facing surfaces. Figure 8 shows the mean deposition velocities of passive horizontal upward-facing surfaces for each condition at distances beyond 2.5 m from the breathing simulator. We specifically looked at these distances because they are beyond any current physical distancing recommendation distance^58^ and because in our prior research, we found only a weak variation in deposition velocities with a distance beyond 2.5 m.^20^ Specifically, relative to the BL condition, deposition velocities for the BL+FiltersO and BL+Filters+ESP conditions were reduced by approximately one-third, a statistically significant decrease (p < 0.01 vs. BL and BL+FanO conditions). Mechanistically, this is not surprising, as PAF units presumably lead to the collection of particles within them, which is a clearance mechanism not possible in their absence. Also notable in terms of developing aerosol clearance procedures, we ran the BL+FanO case as a positive control to examine how moving air around in the classroom impacted particle deposition. The deposition velocity of the BL+FanO condition was slightly increased compared with the BL condition. This likely resulted from the increased air movement introduced by the PAF units, which increases deposition via diffusion and inertia.

**Figure 8.**
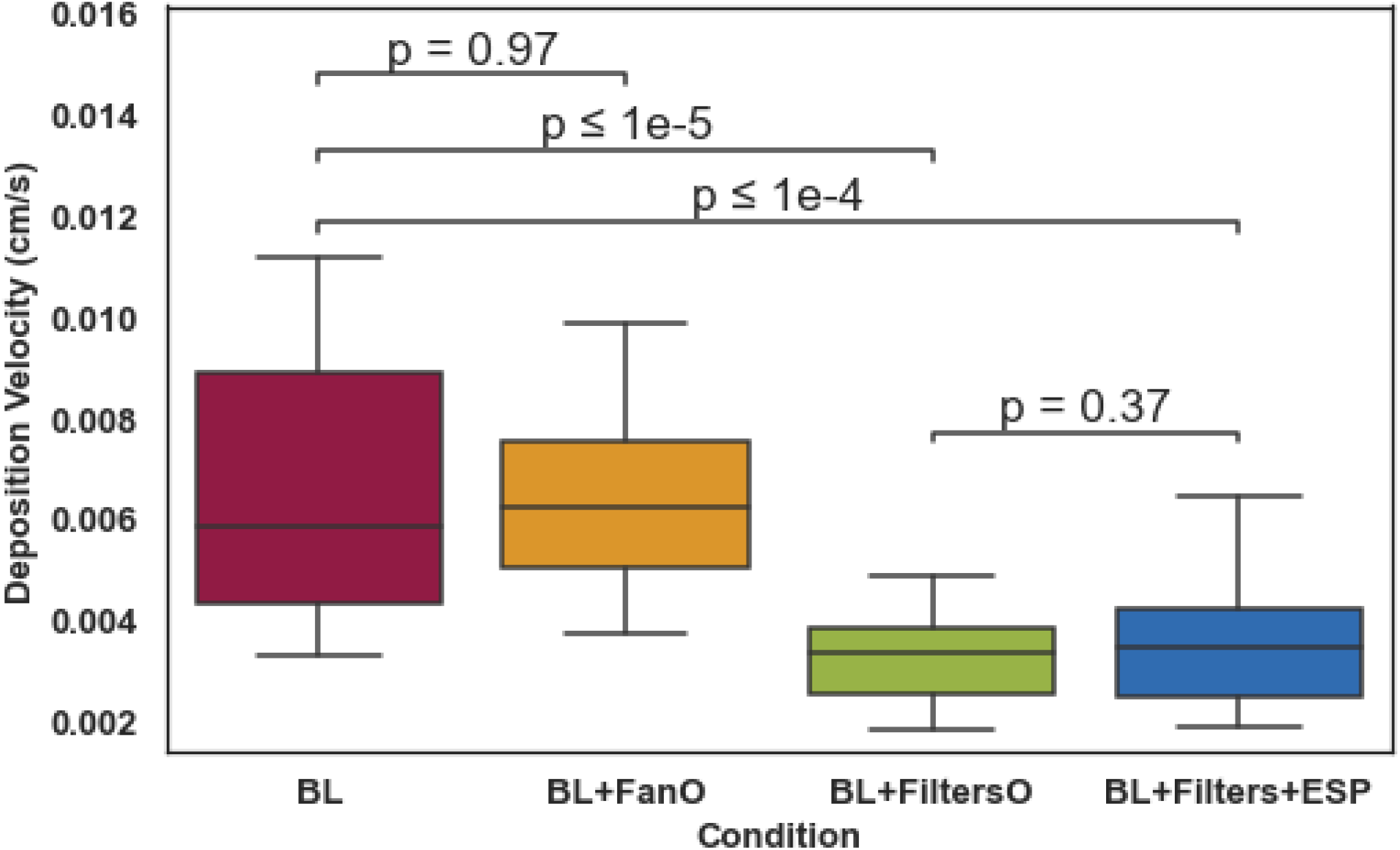
Mean deposition velocities on passive horizontal upward-facing surfaces beyond 2.5 m from the simulator by measurement condition.

Figure 9 shows the deposition flux in the core region of the classroom for each condition. Notably, the scales of these heat maps are not the same, as the peak deposition fluxes of the four conditions differed. That said, BL+FiltersO and BL+Filters+ESP cases share the same scale, and BL and BL+FanO share the same scale. The red dot represents the breathing simulator sitting in the third row, middle seat, with blue boxes representing the location of the PAF units. During all conditions, an area of increased deposition was observed (yellow area) in the ∼1-2 m radius around the breathing simulator, offering some support for the CDC’s physical distancing guidelines.^58^ Outside this zone of higher deposition, deposition flux decreased quickly and was largely uniform in value throughout the rest of the classroom. During the BL+FiltersO and BL+Filters+ESP conditions, higher deposition fluxes were observed on the desk immediately in front of the simulator, CD5-SIM, compared to the BL and BL+FanO conditions. This may have been due to changes in the airflow within the classroom caused by PAF unit operation. Indeed, without the PAF units, most of the exhaled air from the breathing simulator was blown to the right side of the classroom by local airflow but, with the PAF units, the clean recirculated air from the units might have created a stronger zone of stagnant air in front of the simulator. Although the BL+FanO condition altered airflow when running the PAF unit without filters (*i.e.*, fan only), the airflow was much stronger without the filters installed (see Section 1 of supporting information on the supply airflow rate of the PAF unit with and without filters) and may have pushed the zone of stagnant air farther from the desk immediately in front of the breathing simulator.

**Figure 9.**
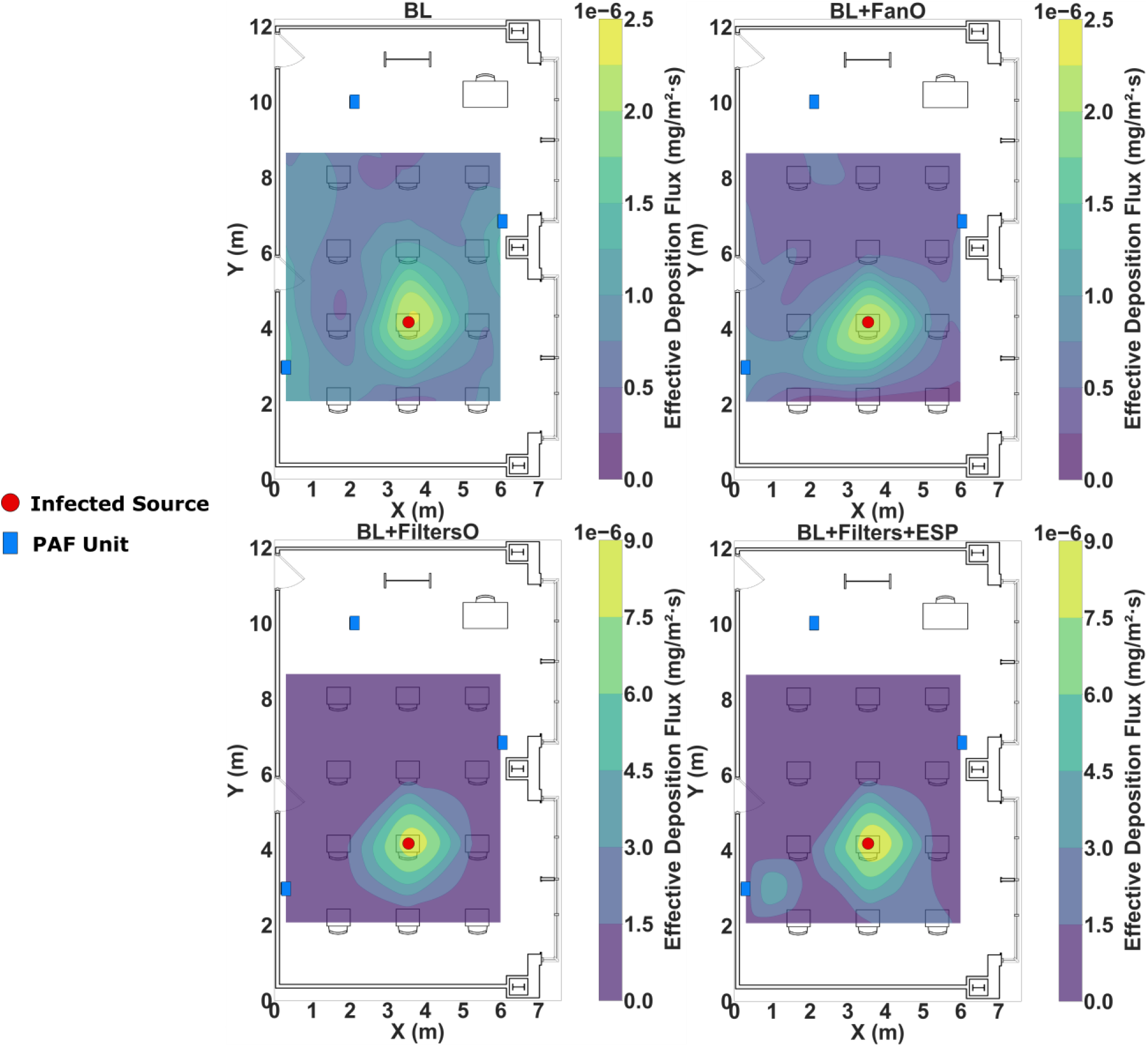
Heat maps of the deposition flux for each of the four measurement conditions.

### 4.2. Local Effectiveness

To quantify the local and whole-room effectiveness of the PAF units, we calculated the local air cleaning effectiveness *η*_*AP*_*i*,*j*_ at location *j* (Figure 10) via the equation:

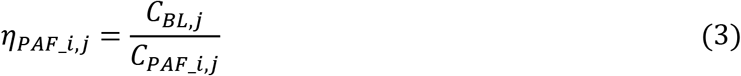

where *C*_*BL,j*_ is the aerosol concentration at location *j* of the BL condition, and *C*_*PAF*_*i*,*j*_ is the aerosol concentration at location *j* of the PAF_*i* condition (*i* = BL+FanO, BL+FiltersO, or BL+Filters+ESP conditions). Values for *η* higher than 1 indicate higher effectiveness at aerosol particle removal versus the BL condition. Box-and-whisker plots resulting from concentration measurements are also shown in Figure 11. Compared to the BL condition, we observed slightly lower particle concentrations throughout the whole classroom for the BL+FiltersO and BL+Filters+ESP conditions (Figure 11; see also Figure 7). However, the measured influence on steady particle concentration is more subtle than measurements of deposition velocity. Greater effectiveness did coincide with the location of the PAF units in relation to areas of greatest airborne particle accumulation. For example, during the BL+FiltersO condition, three zones of greater particle removal relative to the baseline were observed in the top left, bottom left, and middle right (*i.e.*, the three locations of the PAF units). For the BL+Filters+ESP condition, we only observed two zones of greater particle removal vs. BL, middle right and bottom left. The differences in observations may have been due to the change in HVAC operational modes during the time we conducted this study. We conducted this study from April to June 2021 in Rochester, MN. While the flow rate, filtration, temperature, and relative humidity conditions of the classroom were identical across conditions, the BL+FiltersO condition was conducted under a cooling operational mode to maintain these conditions, while BL+Filters+ESP was run under a heating operational mode. Therefore, the more limited impact throughout the whole classroom observed in the BL+Filters+ESP condition may have been due to how the heated air being pumped into the space influenced particle movement, possibly creating a vertical stratification of air that prevented as great of air mixing, relative to the BL+FiltersO condition. The effect of background particles brought in by the HVAC system also presumably affected results as the breathing simulator is a single point of particles. For this reason, we did not see large differences in steady particle concentrations for each of the four measurement conditions. This supports the continued use of CADR or decay-type tests to examine the efficacy of PAF units, while principle steady-tests can yield the same information on CADR difference may be more difficult to discern in such tests.

**Figure 10.**
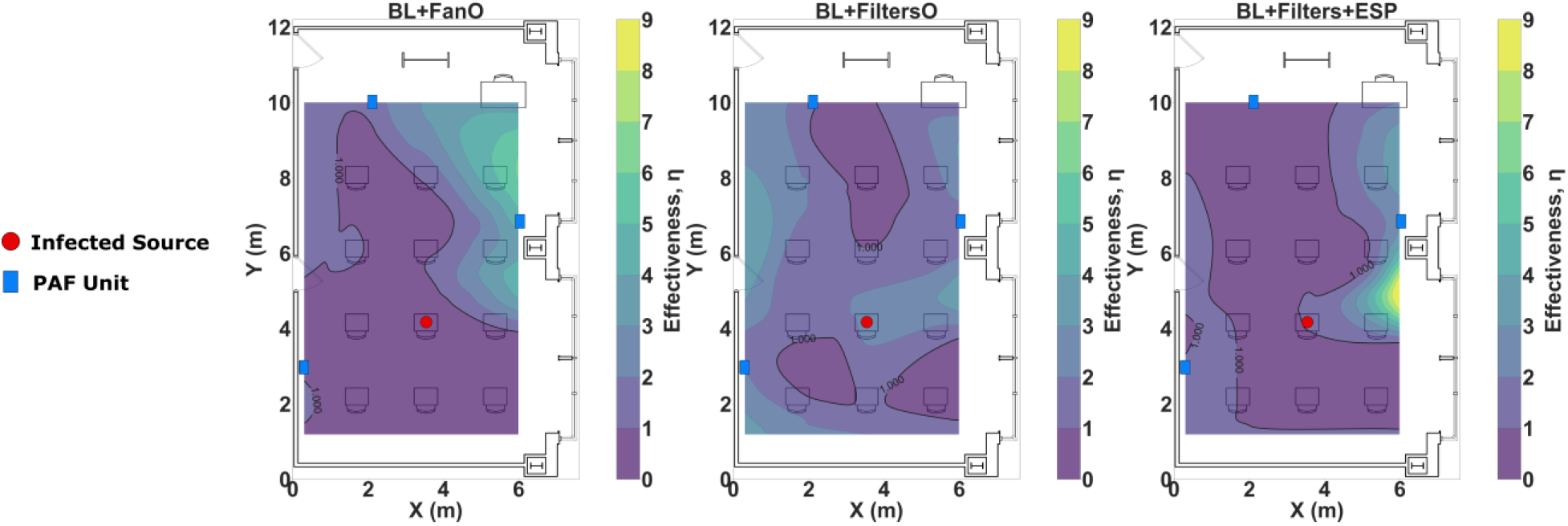
Spatial distribution of particle removal effectiveness for the BL+FanO, BL+FiltersO, and BL+Filters+ESP conditions. All comparisons are relative to BL condition and account for spatial variations in the BL condition.

**Figure 11.**
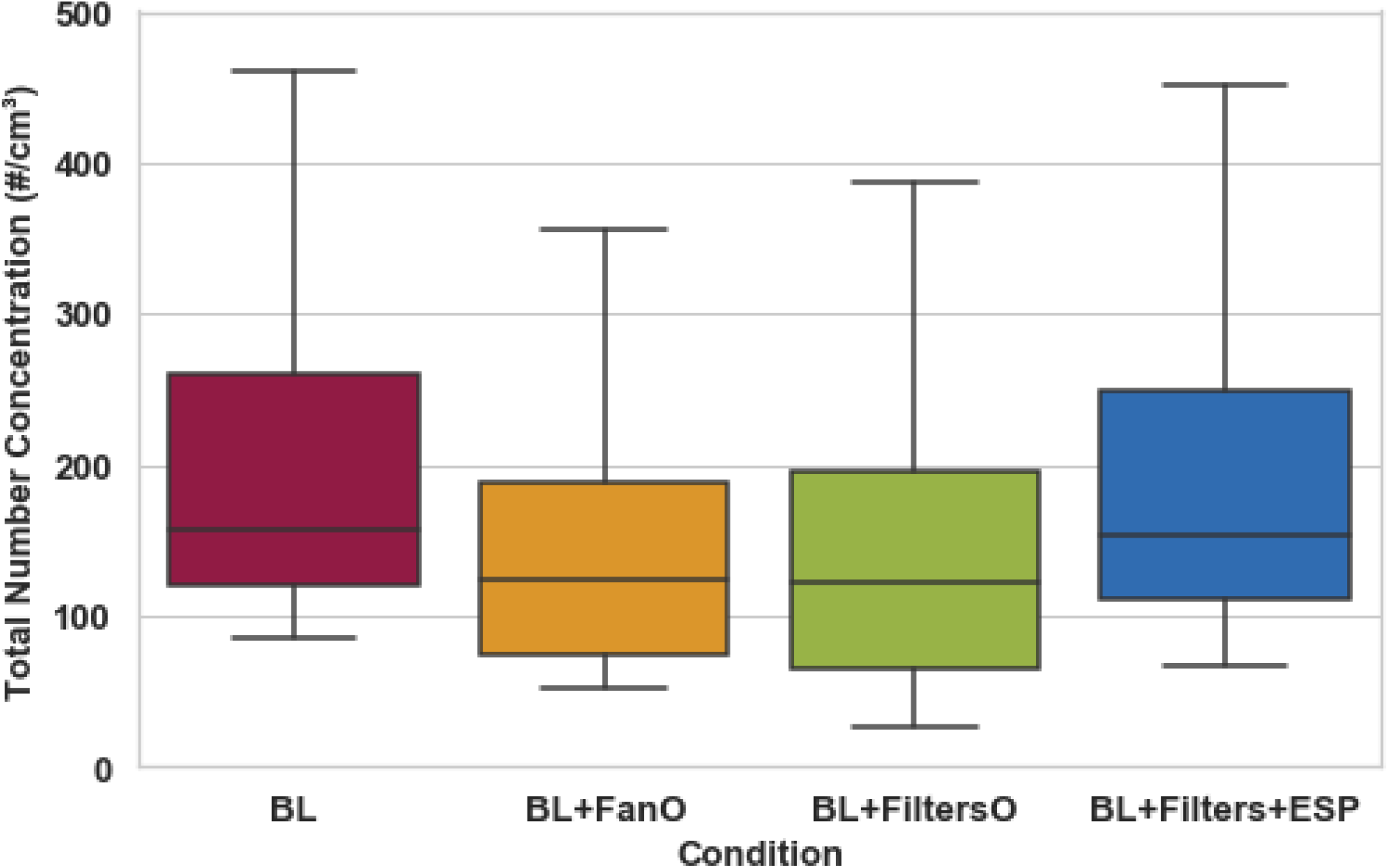
Average total aerosol particle number concentration beyond 2.5 m from simulator by condition.

## 5. Conclusions and Limitations

With in-person instruction in K-12 schools commencing in many districts during the upcoming academic year, it is important to systematically investigate engineering control strategies that may mitigate the infection risks from COVID-19 and other respiratory viruses in schools. This study investigated how PAF units could reduce the localized and whole-room airborne concentration and surface deposition of 1-3 μm fluorescein-tagged aerosol particles injected into the room from a physiologically-correct breathing simulator connected to anatomically-correct respiratory manikin within a mock classroom. Somewhat uniquely, our results address PAF unit influence on both direct aerosol transmission via inhalation and indirect via particle deposition and later transfer, which can occur even for micrometer-scale particles with longer lifetimes in the air. Primary findings are:

- Even for 2 μm particles, the greatest amount of particle deposition was present within ∼2 m of the infectious source. The larger distribution of deposition throughout the rest of the classroom was lower and more uniform when PAF units supplemented the HVAC system versus the HVAC system alone (*i.e.*, the BL condition). Compared with deposition onto passive vertical surfaces, deposition onto passive horizontal upward-facing surfaces was 1-2 orders of magnitude higher because of the effect of gravity on particle behavior. Deposition onto electronic screens and monitors did not show significant differences when paired groupings of electronic devices in the same location were powered on and off, nor did deposition to these surfaces appear different than that to passive horizontal upward-facing surfaces and passive vertical surfaces. These observations are likely due to the weak electrostatic field generated by the screens of these electronic devices. Deposition onto active horizontal downward-facing surfaces (diffusers and return grilles) provided evidence suggestive of how the PAF units during the BL+FiltersO and BL+Filters+ESP were filtering and recirculating clean air back into the space compared to the BL and BL+FanO conditions, as the latter two conditions typically had higher deposition to diffusers and return grilles.
- Compared with the BL condition, the BL+FiltersO and BL+Filters+ESP conditions were observed to have deposition velocities approximately one-third lower outside of 2 m from the infectious source. No differences in deposition velocities were observed between the BL+FiltersO and BL+Filters+ESP conditions, indicating that the ESP likely did not have a marked effect on particle deposition as it is likely not as effective a collection technology as the filter (as suggested in Figure S3 in the supporting information).
- Aerosol particle concentration was greatest at the locations closest to the infectious source, with the distribution of particles around the source affected by the localized airflow. This is congruent with our deposition observations. Also consistent with our deposition observations, the PAF units appeared to reduce the aerosol concentrations throughout most locations in the classroom during the BL+ FiltersO and BL+Filters+ESP conditions. That said, the operational mode of the HVAC system, heating or cooling, appeared to impact how far from each PAF unit increased particle removal effectiveness was observed as these units were supplementing the HVAC system. When the HVAC was in a heating operational mode to maintain our temperature setpoints, the effect of the PAF units was more localized to their location, where the effect was more diffuse when the HVAC was in a cooling operational mode to maintain these setpoints. Nonetheless, PAF units, particularly during the heating season, may better mix a room’s air by minimizing the vertical stratification of air possibly created by warmer air being delivered to the space. This may assist in greater particle removal via the HVAC system and/or the other PAF units in the room.

The limitations of this study include: 1) Aerosol concentrations were measured only at seated breathing height. Concentrations at different heights, especially standing breathing height, may provide more insight as to the dispersion of aerosol particles.; 2) While particle deposition and concentration data were collected at many different locations throughout the classroom, particle lifetime is not directly measured via the methodology employed. This is notable as the lifetime of deposited or suspended particles will determine the viability of the attached virus.; 3) Although the classroom’s environmental conditions were maintained at the same setpoints throughout the whole study, the operational mode of the HVAC system changed with the weather conditions (heating vs. cooling). It caused some inconsistency in the background airflow field.

However, this would be expected in a real-world classroom as the seasons change.; 4) We did not investigate masking. This was an *a priori* choice given the high number of studies that have been completed on mask effectiveness^59^ and the need to detect signals during our deposition experiments; and 5) We did not investigate the precense of other occupants within the classroom. This decision was also made *a priori* to concentrate fully on the impact of PAF units on localized and whole-room deposition and concentration within the classroom. That said, occupants would augment the air flow in the classroom due to movement and thermal plume generation, with increased surface area also contributed by occupant presence, particularly relevant to our deposition measurements.

## Data Availability

The authors confirm that the data supporting the findings of this study are available within the article and its supporting information.

## Acknowledgment

We wish to thank William Baker from Mayo Clinic Respiratory Care for allowing use of the respiratory manikin as well as Eric Heins, Well Living Lab Director of Building Operations, for assisting with continued acquisition of materials for these experiments.

