## Supporting Information for "Localized and Whole-Room Effects of Portable Air Filtration Units on Aerosol Particle Deposition and Concentration in a Classroom Environment"

### 1. Flowrate Measurements of the Recirculating Portable Air Purification (PAF) Unit

The flowrate from the Intellipure Compact PAF unit was measured with and without the filters, as the filters cause a pressure drop and reduce the flow rate provided by the unit's fan. Given that we ran conditions with the filters (BL+FiltersO and BL+Filters+ ESP) and without filters or ESP (BL+FanO), both measurements were important to provide further explanations to the observations made during the study. To perform flowrate measurements, a cardboard box ($28 \times 14 \times 48$ cm$^3$) was attached to the outlet of one of the PAF units to direct and stabilize the air from the outlet. Eight measurement points were distributed evenly over the outlet of the box. The exiting air velocity at each point was measured by a TSI VELOCICALC 9515 probe. Under the with filters and without filters conditions, the overall average exiting air velocity was obtained by averaging the measurements at these eight points. The total air flowrate was calculated by multiplying by the area of the exit of the cardboard box ($0.28 \times 0.14$ m$^2$). We have placed pictures of this setup below in Figure S1a and Figure S1b, and Table S1 shows the data from these measurements.

a. b.
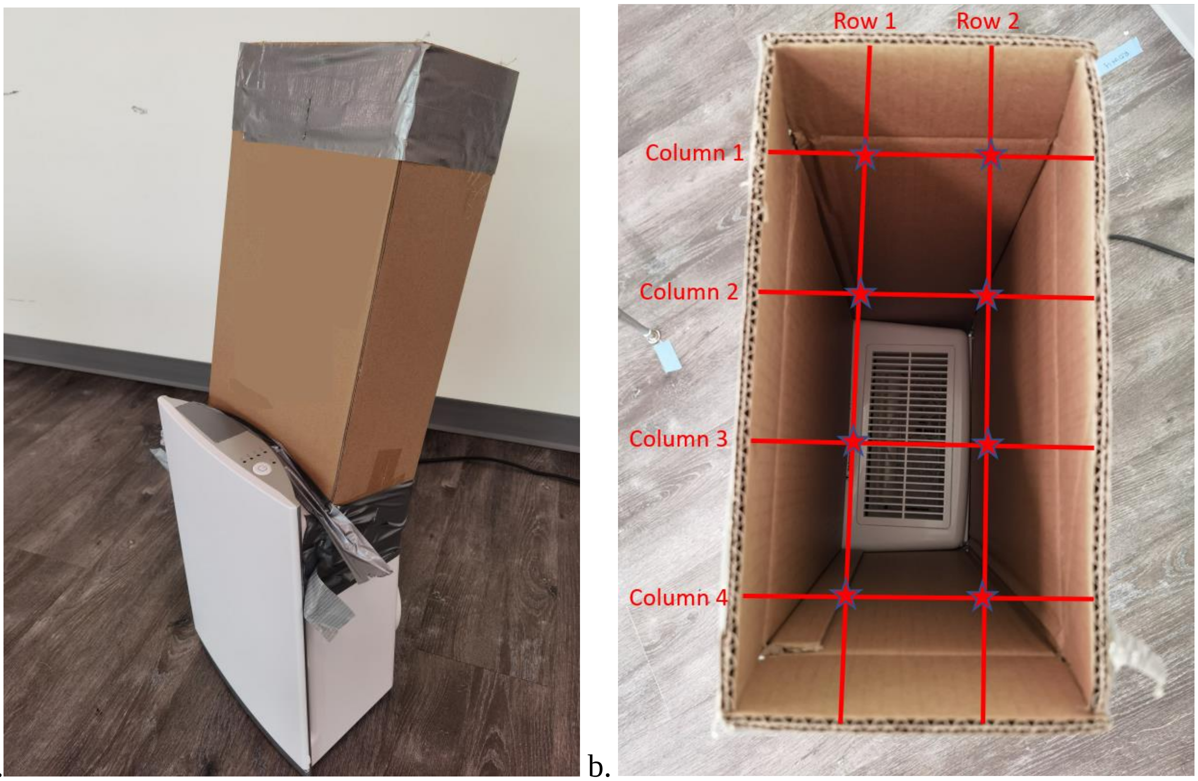


*Figure S1 Flowrate measurement setup (a) and measurement point (b)*

*Table S1 Velocity and flow rate measurement of the air purification unit*

| Condition | Velocity (m/s) | | | | | Flow Rate ($m^3/h$) | Flow Rate (cfm) |
|---|---|---|---|---|---|---|---|
| | | Column 1 | Column 2 | Column 3 | Column 4 | | |
| With Filters | Row 1 | 1.36 | 1.05 | 1.88 | 1.18 | 228 | 134 |
| | Row 2 | 2.92 | 2.13 | 1.38 | 1.10 | | |
| Without Filters | Row 1 | 2.72 | 1.32 | 1.53 | 1.53 | 286 | 168 |
| | Row 2 | 4.56 | 1.92 | 1.28 | 1.36 | | |

### 2. Diffusion and Field Charge Aspects of the Electrostatic Precipitator (ESP) in the PAF Units

We performed additional measurements and calculations to clarify the anticipated extent of particle charging via both diffusion and field charge mechanisms with the ESP section of the PAF units. We first measured the distance from the wires that positively charge particles to the aluminum control grid. Figure S2 provides photographs for where these measurements were made. Then the electric field was estimated as $1.39\text{x}10^5$ V/m with 10kV voltage applied based upon this gap distance. Furthermore, based upon the different manufacturer set device flow rates, we estimated the flow residence time in the particle ionization zone, summarized in Table S2. Note that the "turbo" setting was applied in all reported measurements.

a. b.

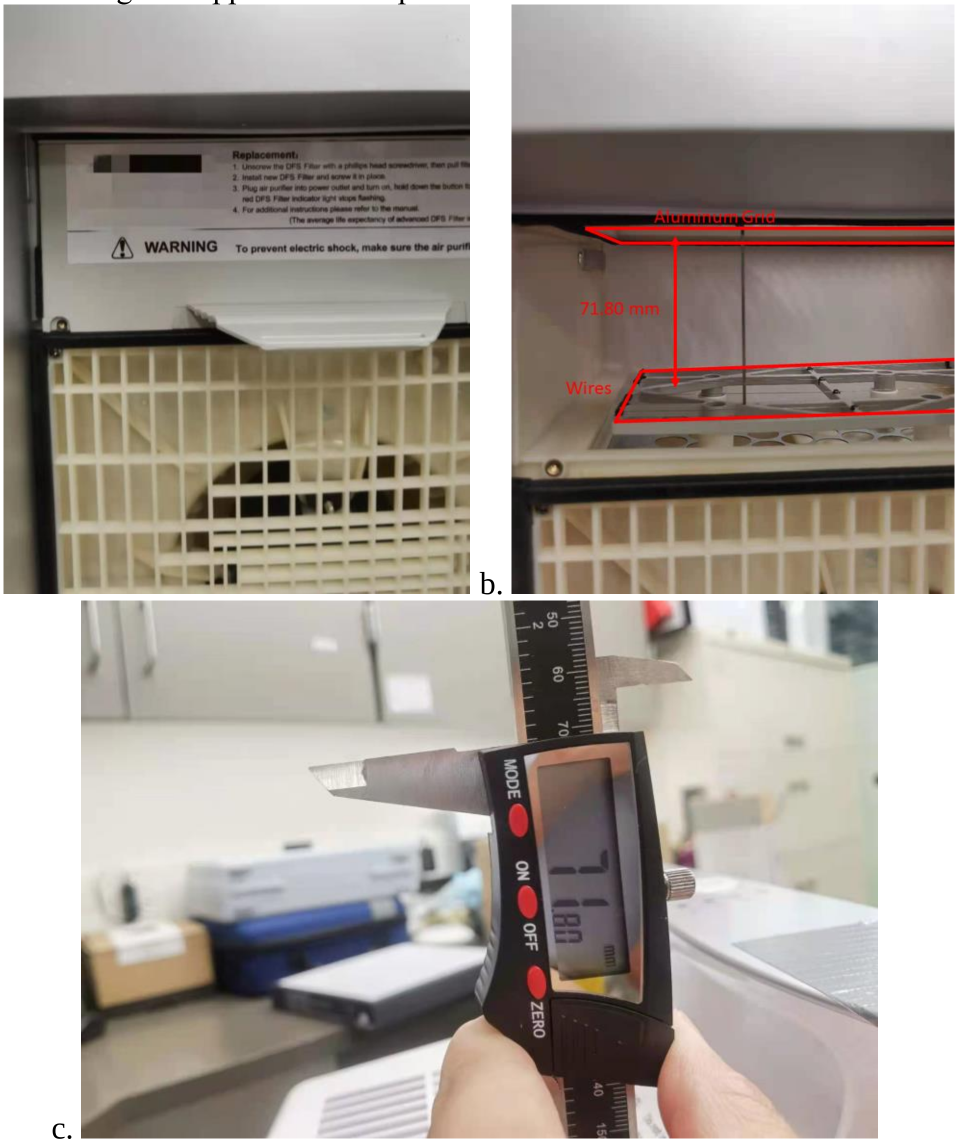


c.

*Figure S2. ESP charge field calculation in PAF units (a. main filter in PAF unit; b. locations of the wires and aluminum grid in PAF unit; c. distance from the grid to the far side of the main filter)*

*Table S2 Residence time calculation under different operation level*

| | Flowrate | Flowrate | Residence Time in ESP field |
|---|---|---|---|
| | CFM | m$^3$/h | Sec |
| **Turbo** | 150 | 256 | 0.032 |
| **High** | 79 | 134 | 0.062 |
| **Medium** | 49 | 83 | 0.099 |
| **Low** | 22 | 37 | 0.2 |

To have a better understanding of the ESP section collection efficiency, we then numerically solved the unipolar charging equations for particles, considering both diffusion and field charging mechanisms. The basic charging equations are:

$$\frac{dn_0}{dt} = -\beta_0 n_0 N_i \ldots (1a)$$

$$\frac{dn_p}{dt} = \beta_{p-1} n_{p-1} N_i - \beta_p n_p N_i \ldots (1b)$$

Here, $n$ denotes the number concentration of aerosol particles; $N_i$ is the ion concentration provided by the manufacturer of 1.75 x 10$^6$ cm$^3$; $\beta_p$ is the collision kernel between particles carrying $p$ charges and ion. An experimentally validated Langevin Dynamics (LD) based model[1,2] of $\beta_p$ was employed for diffusion charging with the consideration of both Coulombic and image force. Conversely, for the field charging mechanism, the charging rate constant for spherical particle of radius $a$ can be expressed by:[3]

$$\beta_p = \frac{eZn_s}{4\varepsilon_0}\left(1 - \frac{n_p}{n_s}\right)^2 \ldots (2a)$$

$$n_s = \{1 + 2[(K-1)/(K+2)]\}\frac{4\pi\varepsilon_0 E_0 a^2}{e} \ldots (2b)$$

where $e$ is the elementary charge, $\varepsilon_0$ is the electric constant, $Z$ is the ion mobility, $n_s$ is the saturation charge of the particle, $E_0$ is the applied electric field, and $K$ is the dielectric constant of the particle. The ion mass and mobility used by Adachi et al.[4] were used for $\beta_p$ calculations (for simplicity, as these are commonly used values). To demonstrate the charging behavior of the particles in the cases of diffusion charging and field charging, the mean particle charge as a function of particle size is plotted in Figure S3a. The corresponding mean electrical mobility ($Z_p$) as a function of charges on the particle ($\bar{z}$) and particle size is shown in Figure S3b. The latter is calculated via the Stokes-Millikan equation:[5]

$$Z_p = \frac{\bar{z}e}{f} \ldots (3a)$$

$$f = \frac{6\pi\mu a}{1 + \frac{\lambda}{a}\left(\gamma_1 + \gamma_2 exp\left(-\frac{\gamma_3 a}{\lambda}\right)\right)} \ldots (3b)$$

With $\lambda$ as the hard sphere mean free path of the gas molecules, $\mu$ as the gas dynamic viscosity, and $\gamma_1 = 1.249, \gamma_2 = 0.4, \gamma_3 = 0.55$ as the empirical constants of the slip correction factor. While technical, both diffusion and field effects act simultaneously on particles, a reasonable approximation is the resulting particle charge is the higher one of the diffusion charging or field charging prediction. Therefore, as is evident in Figure S3, submicrometer particles are likely ionized via the diffusion charging mechanism, while supermicrometer particles are ionized via the field charging mechanism in the ESP of the PAF unit. It is also clearly seen that the number of charges that particles of diameter below 0.3 $\mu m$ acquire is quite limited (less than 1). This is common in ESPs for particles below a critical size: smaller particles require longer residence times or higher ion concentrations to efficiently charge (higher "*nt*" products are needed). The ESP in the PAF unit, therefore, acts predominantly to increase the collection efficiency for micrometer particles.

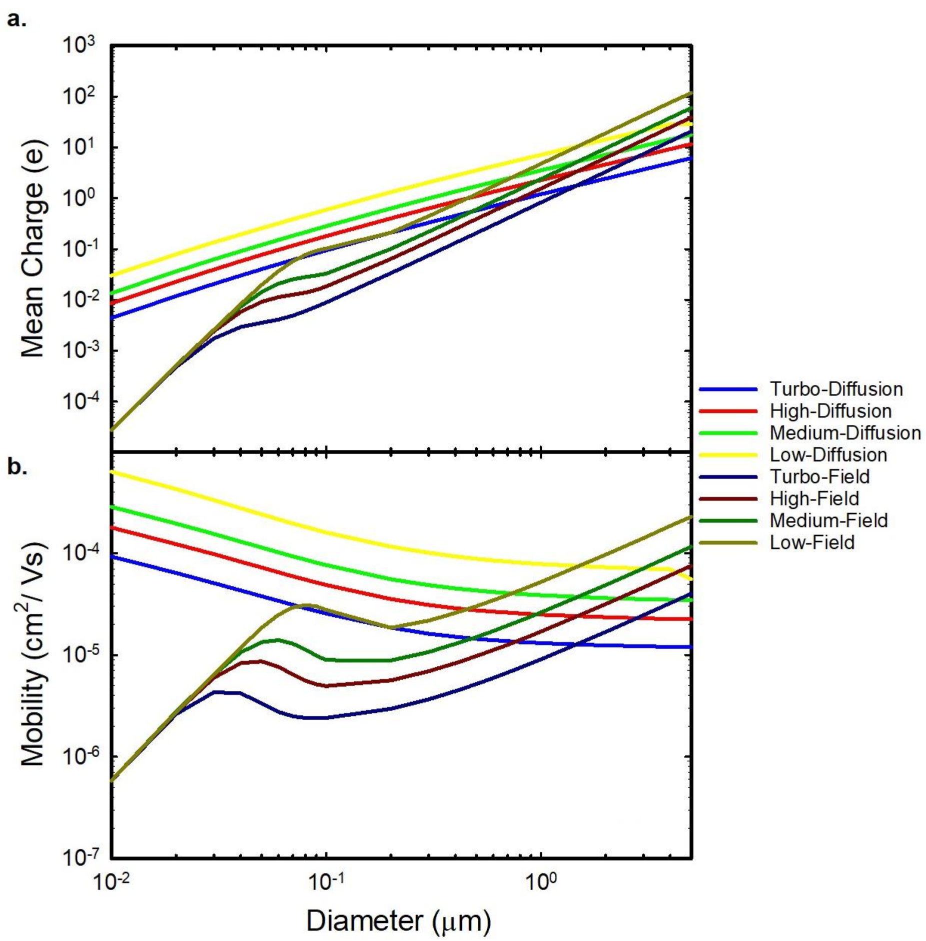


*Figure S3 Comparision of mean particle charge and mean electrical mobility by diffusion charging and field charging mechanism under different operation levels*

### 3. Expanded Aerosol Spatial Distribution Experiment Methodology

This section reviews in more detail the aerosol spatial distribution measurement methodology. Trials were completed with a TSI OPS 3330 placed upon a 32" height cart which was moved every 2 minutes to the locations indicated by the blue squares in Figure S4. The cart had a wire frame that minimally impeded airflow. A reference TSI OPS 3330 was placed on the desk immediately in front of the breathing simulator (CD8).

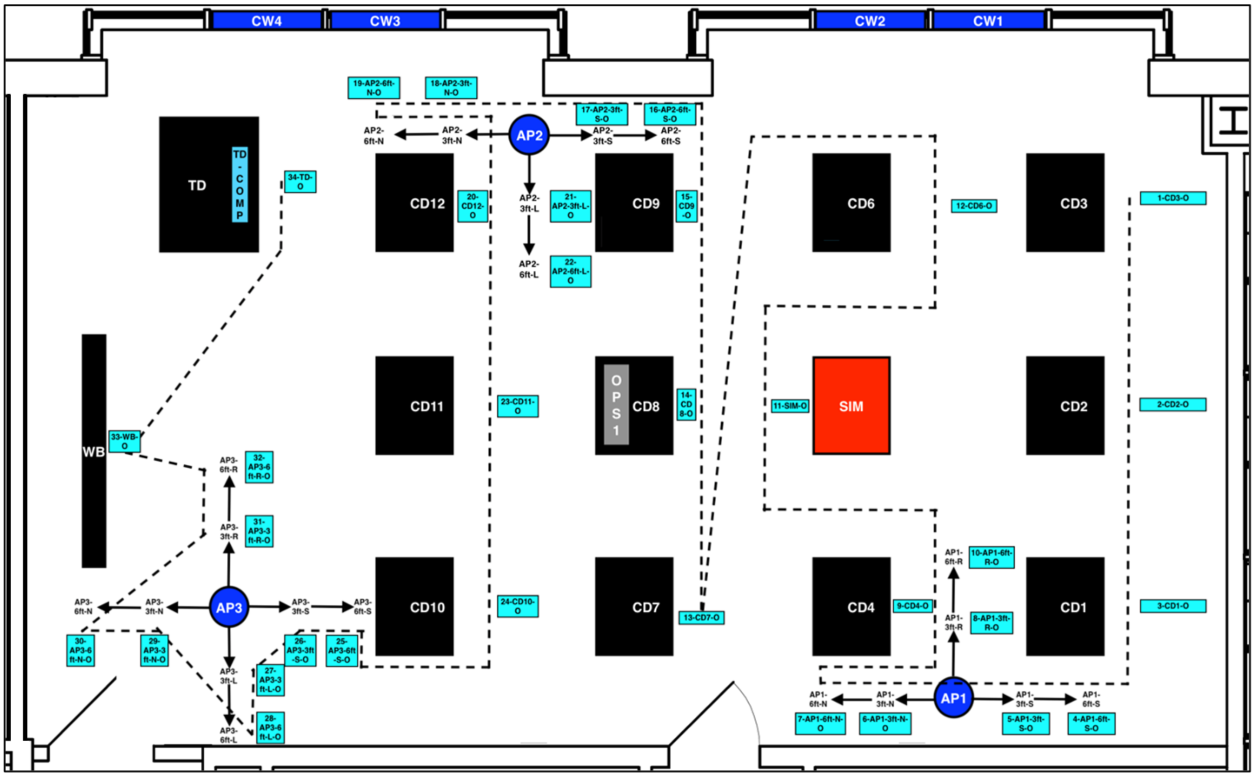


*Figure S4 OPS trial cart moving route*

#### 3.1 Classroom Setup

The classroom was set up identical to that described in the manuscript for the deposition experiments. However, we removed all the shelving units, tall chairs, and vacuum pumps from the space, as these pieces of equipment were unneeded, given that no deposition measurements were being performed during this portion of the study. All the experimental conditions performed were identical to that of the deposition experiments (see Table 1 of the main text).

#### 3.2 Instrument Setup

We brought the two OPS units into the classroom after getting the classroom setup as described above. We placed OPS Unit #2 on the wire cart. We charged OPS Unit #2 the day prior to each experiment, as we used the lithium-ion battery source for this unit to prevent the need to have this unit connected to power throughout the trial when OPS Unit #2 was pushed from location to location. OPS Unit #1 was placed on desk CD8 and was plugged into a power source, with data from this unit used as a marker for the overall room's particle concentration. Both units were run for 3 hours and at a sampling interval of 10 seconds. The wire cart was placed at the initial location on the figure below "1-CD3-O" with OPS Unit #2 sitting atop.

Once the room was set up in the manner described above, we set up the breathing simulator. The setup of the breathing simulator proceeded in the same manner as that described for the deposition experiments above, but we did not place any fluorescein into the solution that we nebulized. Instead, we mixed 20% glycerol with 80% distilled water. OPS data verified that the particle size distribution remained the same as that during the deposition experiments.

#### 3.3 OPS Sampling Procedures

We started both OPS units simultaneously. Once we confirmed both OPS units were running, we allowed them to run for 15 minutes before turning on the simulator to collect background particle concentration data. Following this 15-minute time period, we ran the simulator identically to that described for the deposition experiments. We then exited the room for 60 minutes. At exactly 60 minutes, two Researchers reentered the room. Researcher #1 proceeded to the wire cart where OPS Unit #2 was positioned. Once at the cart, Researcher #1 had Researcher #2, positioned at a set location throughout the duration of the experiment holding a study laptop, note the 'Location Start Time' in a data spreadsheet. 'Location Start Time' came directly from the readout of the OPS Unit #2. Researcher #1 allowed OPS Unit #2 to sample at that location within the classroom for 2 minutes. During those two minutes, Researcher #1 strove to be as still as is reasonably possible while near the wire cart to ensure they minimally impeded airflow from any diffuser, return grille, or PAF unit. At the end of 2 minutes, Researcher #1 had Researcher #2 note the 'Location Stop Time', with the time, again, coming directly from the readout of OPS Unit #2. The same process was repeated at each location, with movement between locations completed methodically, yet expeditiously. The cart was centered over pieces of tape set out on the floor and precisely measured for these experiments. Importantly, the Researchers monitored the breathing simulator throughout the duration of these experiments to ensure that enough solution was still present within the nebulizer and that the simulator is still operating properly. Following the conclusion of sampling at the 34$^{th}$ location, the Researchers shut off the breathing simulator, and exited the room to allow both OPS units to finish their 3-hour run. OPS Unit #2 remained at location "34-TD-O" during this time while OPS Unit #1 remained at CD8. Once the OPS Units finish their 3-hour runs, the Researchers reentered the room and exported the data from both OPS units.

#### 3.4 Before and Between Trials

Before and between trials, the following procedures were completed:

- Thorough cleaning of the classroom to reduce background levels of aerosol particles in the air and to remove particles deposited onto surfaces. All surfaces were cleaned using disinfectant wipes, and the classroom air was scrubbed for more than 24 hours by the room's HVAC system and a standalone commercial PAF unit with a CADR of 1614 $m^3/h$ (950 CFM).
- Replacement of the HEPA capsule connected to the mass flow controller in the breathing simulator, with the cleaned and dried hose barb connections wrapped with plumber's tape to prevent leaks placed onto a new HEPA capsule.

- A check that the battery for OPS Unit #2 was charged. This was critical for the reasons mentioned above.
